# Shared genetic architecture of brain age gap across 30 cohorts worldwide

**DOI:** 10.64898/2025.12.23.25342890

**Authors:** Vilte Baltramonaityte, Philippe Jawinski, Marlene Staginnus, Mina Shahisavandi, Boglarka Z Kovacs, Isabel K Schuurmans, Constantinos Constantinides, Ahmad R Hariri, Alexander Teumer, Amanda L Rodrigue, Ami Tsuchida, Amirhossein Manzouri, Andriana Karuk, Anna E Fürtjes, Annalisa Lella, Annchen R Knodt, Antonia Jüllig, Avshalom Caspi, Benedicto Crespo-Facorro, Brenda WJH Penninx, Catharina Lavebratt, Christine Lochner, Clarissa L Yasuda, Dag Alnæs, Dan J Stein, Daniel H Mathalon, David C Glahn, Dennis Klose, Dylan J Kiltschewskij, Edith Pomarol-Clotet, Estela M Bruxel, Fabrice Crivello, Fernando Cendes, Gail Davies, Giovanni A Salum, Hans J Grabe, Heather J Zar, Henning Tiemeier, Henry Völzke, Hervé Lemaître, Iscia Lopes-Cendes, Ítalo Karmann Aventurato, Jean Shin, Jessica A. Turner, Joanna M Wardlaw, John Blangero, Jonathan C Ipser, Judith M Ford, Kang Sim, Karen Sugden, Katharina Wittfeld, Kristina Salontaji, Kristoffer NT Månsson, L. Elliot Hong, Lars T Westlye, Lianne Schmaal, Lucas T Ito, Lucas Scárdua-Silva, Marcos L Santoro, María Alemany-Navarro, Mark E Bastin, Mary S Mufford, Melissa J Green, Murray J Cairns, Nadine Parker, Nathaniel W. McGregor, Ole A Andreassen, Oliver Gruber, Oliver J Watkeys, Pedro M Pan, Peter Kochunov, Qian Hui Chew, Rafael Romero-Garcia, Raymond Salvador, Reremoana Theodore, Richie Poulton, Robin Bülow, Rodrigo A Bressan, Ryan L Muetzel, Sebastian Markett, Serena Defina, Sheri-Michelle Koopowitz, Shir Dahan, Simon R Cox, Sintia Belangero, So M Vijayakumar Kamalakannan, Sophia I Thomopoulos, Stefan Frenzel, Terrie E Moffitt, Theo GM van Erp, Tomas Furmark, Tomas Paus, Uwe Völker, Vince D. Calhoun, Yann Quidé, Younghwa Lee, Yuri Milaneschi, Zdenka Pausova, Paul M Thompson, Laura KM Han, Jean-Baptiste Pingault, James H Cole, Charlotte AM Cecil, Sarah E Medland, Danai Dima, Esther Walton

**Affiliations:** Department of Psychology, University of Bath, Bath, United Kingdom; Department of Psychology, Humboldt-Universität zu Berlin, Berlin, Germany; The Generation R Study Group, Erasmus University Medical Center, Rotterdam, the Netherlands; Department of Child and Adolescent Psychiatry/Psychology, Erasmus Medical Center Rotterdam, Rotterdam, The Netherlands; Department of Psychology & Neuroscience, Duke University, Durham, United States; Department of Psychiatry and Psychotherapy, University Medicine Greifswald, Greifswald, Germany; German Center of Neurodegenerative Diseases (DZNE), Site Rostock/Greifswald, Greifswald, Germany; Department of Psychiatry and Behavioral Sciences, Boston Children’s Hospital, Boston, United States; Department of Psychiatry, Harvard Medical School, Boston, United States; Centre for Psychiatry Research, Department of Clinical Neuroscience, Karolinska Institutet, & Stockholm Health Care Services, Stockholm, Sweden; Department of Psychology, Stockholm University, Stockholm, Sweden; Department of Clinical Psychology and Psychotherapy, Babeș-Bolyai University, Cluj-Napoca, Romania; FIDMAG Germanes Hospitalàries Research Foundation, Barcelona, Spain; Centro de Investigación Biomédica en Red de Salud Mental (CIBERSAM), Instituto de Salud Carlos III, Madrid, Spain; Lothian Birth Cohorts, Department of Psychology, University of Edinburgh, Edinburgh, United Kingdom; Lothian Birth Cohorts, Edinburgh Futures Institute, University of Edinburgh, Edinburgh, United Kingdom; Department of Experimental Psychology and Cognition Research, Justus-Liebig-University, Gießen, Germany; Institute of Psychiatry, Psychology, & Neuroscience, King’s College London, London, England; Hospital Universitario Virgen del Rocio, IBIS-CSIC, CIBERSAM, Sevilla, Spain; Department of Psychiatry, Amsterdam UMC, Vrije Universiteit Amsterdam, Amsterdam, The Netherlands; Molecular Medicine and Surgery, Karolinska Institutet, Stockholm, Sweden; Center for Molecular Medicine, Stockholm, Sweden; SAMRC Unit on Risk and Resilience in Mental Disorders, Department of Psychiatry, Stellenbosch University, Tygerberg, South Africa; Department of Neurology, State University of Campinas, Campinas, Brazil; Brazilian Institute of Neuroscience and Neurotechnology, Brazil; Department of Psychology, University of Oslo, Oslo, Norway; Center for Precision Psychiatry, Division of Mental Health and Addiction, University of Oslo and Oslo University Hospital, Oslo, Norway; South Africa Medical Research Council Unit on Child and Adolescent Health, University of Cape Town, Cape Town, South Africa; Department of Psychiatry and Behavioral Sciences, University of California, San Francisco, California, United States; School of Biomedical Sciences and Pharmacy, The University of Newcastle, Newcastle, Australia; Precision Medicine Research Program, Hunter Medical Research Institute, Newcastle, Australia; Department of Medical Genetics and Genomic Medicine, School of Medical Sciences, University of Campinas (UNICAMP), Campinas, Brazil; GIN, IMN UMR 5293, CEA, CNRS, Université de Bordeaux, Bordeaux, France; Department of Psychiatry and Legal Medicine, Universidade Federal do Rio Grande do Sul, Porto Alegre, Brazil; Child Mind Institute, New York, United States; Department of Paediatrics and Child Health, Red Cross War Memorial Children’s Hospital, Cape Town, South Africa; Harvard T.H. Chan School of Public Health, Department of Behavioral and Social Sciences, Boston, United States; University Medicine Greifswald, Institute for Community Medicine, Greifswald, Germany; Centre de recherche CHU Sainte-Justine and University of Montreal Montréal Canada; Department of Psychiatry and Behavioral Health, Wexner Medical Center, Ohio State University, Columbus, United States; Institute of Neuroscience and Cardiovascular Research, University of Edinburgh, Edinburgh, United Kingdom; UK Dementia Research Institute Centre, University of Edinburgh, Edinburgh, United Kingdom; Department of Psychiatry and Mental Health, Neuroscience Institute, University of Cape Town, Cape Town, South Africa; Dept of Veterans Affairs, San Francisco, California, United States; West Region, Institute of Mental Health, Singapore, Singapore; Department of Psychiatry, The University of Texas Health Science Center, Houston, United States; Centre for Youth Mental Health, University of Melbourne, Melbourne, Australia; Discipline of Molecular Biology - Department of Biochemistry, Universidade Federal de São Paulo (UNIFESP), São Paulo, Brazil; Translational Psychiatry Group, Instituto de Biomedicina de Sevilla (IBiS)-CSIC, Seville, Spain; Centre for Clinical Brain Sciences, University of Edinburgh, Edinburgh, United Kingdom; Division of Human Genetics, University of the Witwatersrand, Johannesburg, South Africa; School of Clinical Medicine, University of New South Wales, Sydney, Australia; Section for Experimental Psychopathology and Neuroimaging, Department of General Psychiatry, Heidelberg University, Heidelberg, Germany; School of Clinical Medicine, Discipline of Psychiatry and Mental Health, University of New South Wales, Randwick, Australia; Laboratory of Integrative Neurosciences (LiNC), Universidade Federal de São Paulo (UNIFESP), São Paulo, Brazil; Department of Medical Physiology and Biophysics, Seville, Spain; Instituto de Biomedicina de Sevilla (IBiS) HUVR/CSIC, CIBERSAM, ISCIII, Seville, Span; Dunedin Multidisciplinary Health and Development Research Unit, University of Otago, Dunedin, New Zealand; Institute for Diagnostic Radiology and Neuroradiology, Greifswald, Germany; Department of Radiology and Nuclear Medicine, Erasmus University Medical Center, Rotterdam, The Netherlands; Clinical Neuroimaging Laboratory, Galway Neuroscience Center, College of Medicine, Nursing & Health Sciences, University of Galway, Galway, Ireland; Department of Psychiatry, Universidade Federal de São Paulo, São Paulo, Brazil; Graduate Program in Structural and Functional Biology, Universidade Federal de São Paulo, São Paulo, Brazil; Imaging Genetics Center, Mark and Mary Stevens Neuroimaging and Informatics Institute, Keck School of Medicine, University of Southern California, Marina del Rey, United States; Clinical Translational Neuroscience Laboratory, Department of Psychiatry and Human Behavior, University of California Irvine, Irvine, United States; Center for the Neurobiology of Learning and Memory, University of California Irvine, Irvine, United States; Department of Psychology, Uppsala University, Uppsala, Sweden; Departments of Psychiatry and Neuroscience, University of Montreal, Montreal, Canada; Department Functional Genomics, University Medicine Greifswald, Greifswald, Germany; Tri-institutional Center for Translational Research in Neuroimaging and Data Science (TReNDS), Georgia State, Georgia Tech, Emory, Atlanta, United States; NeuroRecovery Research hub, School of Psychology, UNSW Sydney, Sydney, Australia; Centre for Pain IMPACT, Neuroscience Research Australia, Randwick, Australia; Mental Health Program, Amsterdam Public Health, Amsterdam, The Netherlands; Complex Trait Genetics program, Amsterdam Neuroscience, Amsterdam, The Netherlands; Department of Clinical, Educational and Health Psychology London, University College London, United Kingdom; Hawkes Institute, Department of Computer Science, University College London, London, United Kingdom; Psychiatric Genetics, QIMR Berghofer Medical Research Institute, Brisbane, Australia; School of Psychology, University of Queensland, Brisbane, Australia; Department of Psychology and Neuroscience, City St George’s, University of London, London, United Kingdom; Department of Neuroimaging, Institute of Psychiatry, Psychology and Neuroscience, King’s College London, London, United Kingdom; Orygen, The National Centre of Excellence in Youth Mental Health, Parkville, Australia; South Africa Medical Research Council Unit on Risk and Resilience in Mental Disorders, Department of Psychiatry, University of Cape Town, Cape Town, South Africa; DZHK (German Center for Cardiovascular Research), Partner Site Greifswald, Greifswald, Germany; Amsterdam Neuroscience, Mood, Anxiety, Psychosis, Sleep & Stress program, Amsterdam, The Netherlands; Amsterdam Public Health, Mental Health program, Amsterdam, The Netherlands; Human and Systems Genetics Working Group, Department of Genetics, Stellenbosch University, Stellenbosch, South Africa; School of Philosophy, Psychology & Language Sciences, The University of Edinburgh, Edinburgh, United Kingdom; Department of Human Genetics and South Texas Diabetes and Obesity Institute, School of Medicine, University of Texas of the Rio Grande Valley, Brownsville, TX, United States

## Abstract

Deviations from normative brain ageing trajectories are linked to a wide range of adverse health outcomes. A number of brain age prediction models have been developed, based on various neuroimaging modalities, machine learning algorithms, training samples, and age ranges. However, it remains unknown whether these models converge on a shared genetic liability, and whether capturing this shared signal could provide a more sensitive marker of brain health than any single model alone. We first conducted a new brain age gap (BAG) GWAS in a sample of 60,735 individuals across 29 cohorts worldwide, and then applied genomic structural equation modelling to examine the shared genetic variance between five prior BAG GWASs and our new analysis, using a single latent BAG factor (30 cohorts overall). All six BAG GWASs loaded onto a single factor, explaining 63% of the total genetic variance. We identified 19 independent SNPs associated with the BAG factor, including four novel associations. Genetically, the BAG factor was positively correlated with multiple traits, with blood pressure, smoking, longevity, autism, and sleep showing putatively causal effects. A polygenic score (PGS) for the BAG factor showed associations with phenotypic BAGs already in childhood, with stronger links observed in adulthood. Phenome-wide association analyses indicated that BAG factor PGS captured associations with more health traits than individual BAG PGSs. Our findings underscore the importance of considering the shared variance across different BAG constructs to identify robust correlates of poor brain health.

## Introduction

As we age, the brain develops and changes with considerable variability across individuals (Raz & Rodrigue, 2006). Such deviations from a typical brain ageing trajectory are important correlates of poor health outcomes (Cole et al., 2018). Brain age gap (BAG) – conceptualised as the difference between chronological and brain predicted age (e.g., from brain scans) – has become a valuable neuroimaging marker within lifespan neuroscience. BAG has recently been linked to over 200 traits, including disease risk factors (e.g., increased diastolic blood pressure and body mass index), cognitive function, and non-communicable diseases (e.g., type I and type II diabetes) (Jawinski et al., 2025; Kolbeinsson et al., 2020).

In recent years, a growing number of brain age prediction models have been developed. Some models have been used in genome-wide association studies (GWASs) to characterise the genetic underpinnings of BAG, showing single-nucleotide polymorphism (SNP)-based heritability estimates ranging from 0.23 to 0.47 (Jawinski et al., 2025; Leonardsen et al., 2023; Smith et al., 2020; Wen et al., 2024), which are generally higher than estimates for psychiatric disorders (Baselmans et al., 2021). Furthermore, in line with phenotypic research, brain-age GWASs identified potential causal effects of BAG on health outcomes, such as blood pressure and diabetes (Jawinski et al., 2025). Understanding the genetic influence on BAG may therefore help identify risk factors for poorer brain health in later life and shed light on the biological mechanisms underpinning variability in the brain ageing process.

Despite advances in this field, it is currently unclear to what extent different brain age models capture a shared underlying genetic signal. These models have been based on various neuroimaging modalities (T1-weighted structural magnetic resonance imaging [MRI], diffusion imaging, and functional MRI), machine learning algorithms, training samples, and age ranges (Soumya Kumari & Sundarrajan, 2024). Observational studies demonstrate that BAG predictions from distinct models do not always correlate highly with each other (e.g., correlations between BAGs: *r* = 0.45-0.64; Bacas et al., 2023), which may be partly due to these heterogeneous methodological choices, as well as training population differences (e.g., in age, socio-economic background, imaging acquisition protocols) and measurement error. Modelling the joint genetic architecture of different brain age GWASs would enable us to extract a latent BAG factor, allowing us to (1) assess the degree to which genetic associations with BAG are shared across multiple models, (2) boost power for genetic variant discovery, and (3) reduce measurement error. True brain age values can only ever be approximated and depend on the accuracy of the underlying age-prediction models. Low-accuracy models may be too lenient, while highly accurate models may remove biologically meaningful variance or reflect overfitting, thereby limiting the ability to capture clinically or genetically relevant effects (Bashyam et al., 2020; Schulz et al., 2025). To extract the relevant biological signal and reduce measurement noise, we need to aggregate shared genetic variance across different BAG models.

While previous BAG GWASs were based on models with high predictive accuracy (mean absolute error of ∼3 years), they were all based on the UK Biobank (UKBB) cohort, thereby limiting global representation. The Enhancing Neuro Imaging Genetics Through Meta-Analyses (ENIGMA; Thompson et al., 2020). Consortium has pioneered a global approach to neuroimaging genetics and has developed a brain age model that is based on standard FreeSurfer output and is therefore easily scalable across cohorts worldwide (Han et al., 2021). We thus first conducted a large BAG GWAS based on FreeSurfer-derived imaging features, enabling us to combine data across 29 cohorts worldwide and extending beyond the current focus on the UKBB. Subsequently, we applied genomic structural equation modelling (Genomic SEM; Grotzinger et al., 2019) to examine the shared genetic variance across six brain age GWASs, including the present study, using a single latent BAG factor. A latent factor approach may provide a more accurate measure of BAG, capturing shared variance across different imaging-derived predictors (e.g., grey matter volume, thickness, surface area) and thus providing less biased estimates of BAG. We then conducted a GWAS on this latent factor, followed by a comprehensive set of analyses to investigate the biological pathways, shared genetic influences, causal relationships, polygenic risk, and phenome-wide associations with BAG. To our knowledge, this is the first study to integrate multiple BAG GWASs into a unified latent genetic factor, enabling a more robust investigation of its biological and clinical relevance.

## Method

### ENIGMA GWAS

As prior BAG GWASs were primarily based on the UKBB, we embarked on a large-scale international analysis that integrated genomics and standard FreeSurfer output from 29 cohorts worldwide (see map; *N*=60,735). Structural T1-weighted scans were processed using FreeSurfer (Fischl, 2012) resulting in a total of 77 features based on the Desikan/Killiany atlas (Desikan et al., 2006). The 77 features were used to derive BAG as detailed in Han et al. (2021). We first conducted GWASs on the ENIGMA model (BAG_Han_) at the cohort level, and then meta-analysed the results using METAL. For further details see **Supplementary Material (SM) Methods section 1.1** and **Table 1**. Descriptive cohort characteristics, as well as imaging and genotype details, can be found in **Tables S1-S4**.

**Table 1.** Six studies contributing to the genomic SEM GWAS of brain age gap factor.

| Model Name | GWAS |  |  |  |  |  |  |  | Brain age derivation |  |  |  |  |  | Reference |
| --- | --- | --- | --- | --- | --- | --- | --- | --- | --- | --- | --- | --- | --- | --- | --- |
| | *N* | Age range | % females | Ethnicity | Sample | MAE (years) | $h^2$ (SE) | Loci | Imaging modality | Algorithm | Sample | Training age range | MAE (years) | *r* | |
| BAG<sub>Han</sub> (*ENIGMA*) | 60,735 (N<sub>UKBB</sub>=46,322; N<sub>ENIGMA</sub>=14,413) | 18-75 | 53.7% | EUR primarily | 29 datasets (inc. UKBB) | 9.4 | 0.21 (0.01) | 29 | T1w MRI | Ridge regression | 19 datasets | 18-75 | male=6.5<br>female=6.8 | female=0.83<br>male=0.85 | Han et al. (2021); and the present study |
| BAG<sub>Leonardsen</sub> (*pyment*) | 28,104 | 40-84y | 51.6% | EUR | UKBB | 2.45 | 0.27 (0.04) | 8 | T1w MRI | SFCN-reg | 21 datasets (incl. UKBB) | 3-95y | 2.47 | 0.975 | Leonardsen et al. (2023) |
| BAG<sub>Wen</sub> (*brain BAG*) | 30,108 | Approx. 40-84y | NA | EUR | UKBB | NA | 0.47 (0.02) | 11 | Multimodal MRI (T1w, dMRI, rs-fMRI) | Support vector machine regression | UKBB | 46-82y | female=3.5<br>male = 3.7 | female=0.79<br>male=0.80 | Wen et al. (2024); Tian et al. (2023) |
| BAG<sub>Smith</sub> (*62 modes*) | 10,612 | Approx. 40-84y | ~54% | EUR | UKBB | 2.9 | 0.23 (0.05) | 1 | Multimodal MRI (T1w, dMRI, rs-fMRI) | Independent component analysis; regression-based models | UKBB | 45-80y | 2.9 | NA | Smith et al. (2020) |
| BAG<sub>Kaufmann</sub> (*brainage*) | 20,170 | 40-70* | 53%* | EUR | UKBB | NA | 0.24 (0.03) | 1 | T1w MRI | Gradient tree boosting with XGBoost | 41 datasets (inc. UKBB) | 3-89y | 4.78* | female=0.93<br>male=0.94 | Kaufmann et al. (2019) |
| BAG<sub>Jawinski</sub> (*combined BAG*) | 52,890<sup>1</sup> | 45-82 | 52.2% | EUR | UKBB and LIFE-Adult | 3.09 | 0.28 (0.02) | 39 | T1w MRI | Relevance vector machine and XGBoost | UKBB | 45-82y | 3.09 | 0.86 | Jawinski et al. (2025) |

*Note.* For BAG<sub>Leonardsen</sub> we used the regression model (SFCN-reg). For BAG<sub>Wen</sub> we used the multimodal ‘Brain PhenoBAG’ derived from grey matter, white matter, and functional connectivity metrics. For BAG<sub>Smith</sub> we used the all-in-one model incorporating deltas from all 62 modes in a single model (code: V0140). For BAG<sub>Jawinski</sub> we used the combined grey and white matter model. T1w MRI = T1-weighted magnetic resonance imaging; dMRI = diffusion magnetic resonance imaging; rs-fMRI = resting-state functional magnetic resonance imaging; BAG = brain age gap; EUR = European; GWAS = genome-wide association study; *N* = sample size; Loci = genome-wide significant SNPs clumped into approximately independent regions based on linkage disequilibrium thresholds and genomic distance, as reported in the original publications; MAE = mean absolute error; NA = not available; SNPs = single nucleotide polymorphisms; UKBB = UK Biobank; $h^2$ = SNP-based heritability; Combined BAG = combined grey and white matter BAG. <sup>1</sup> GWAS originally conducted with 54,890 individuals; re-run with 52,890 to hold out 2,000 for
polygenic score analyses. \*No mean absolute error (MAE) reported in the original publication. An independent study on a separate dataset by Blake (2021) identified a MAE of 4.78. All BAG GWASs controlled for age, sex, and genetic PCs. Other covariates were study specific. For example, total intracranial volume was controlled for in BAG<sub>Han</sub>, BAG<sub>Wen</sub>, and BAG<sub>Jawinski</sub> only. Genomic SEM = genomic structural equation modelling.

### Genetic data sources for brain age gap

In addition to the GWAS conducted as part of the present study, we also selected five additional BAG GWASs with publicly available genetic summary statistics from previously published studies by Leonardsen et al. (2023), Wen et al. (2024), Smith et al. (2020), Kaufmann et al. (2019), and Jawinski et al. (2025). Throughout the manuscript, we refer to these models as BAG_author.surname_. The models differed in terms of imaging modality, input features, machine learning algorithms, samples, and age ranges. BAG was computed as the difference between predicted brain age and chronological age. A descriptive overview of the six models is provided in **Table 1**. A schematic summary of the analysis workflow is shown in **Figure 1**.

**Figure 1.**
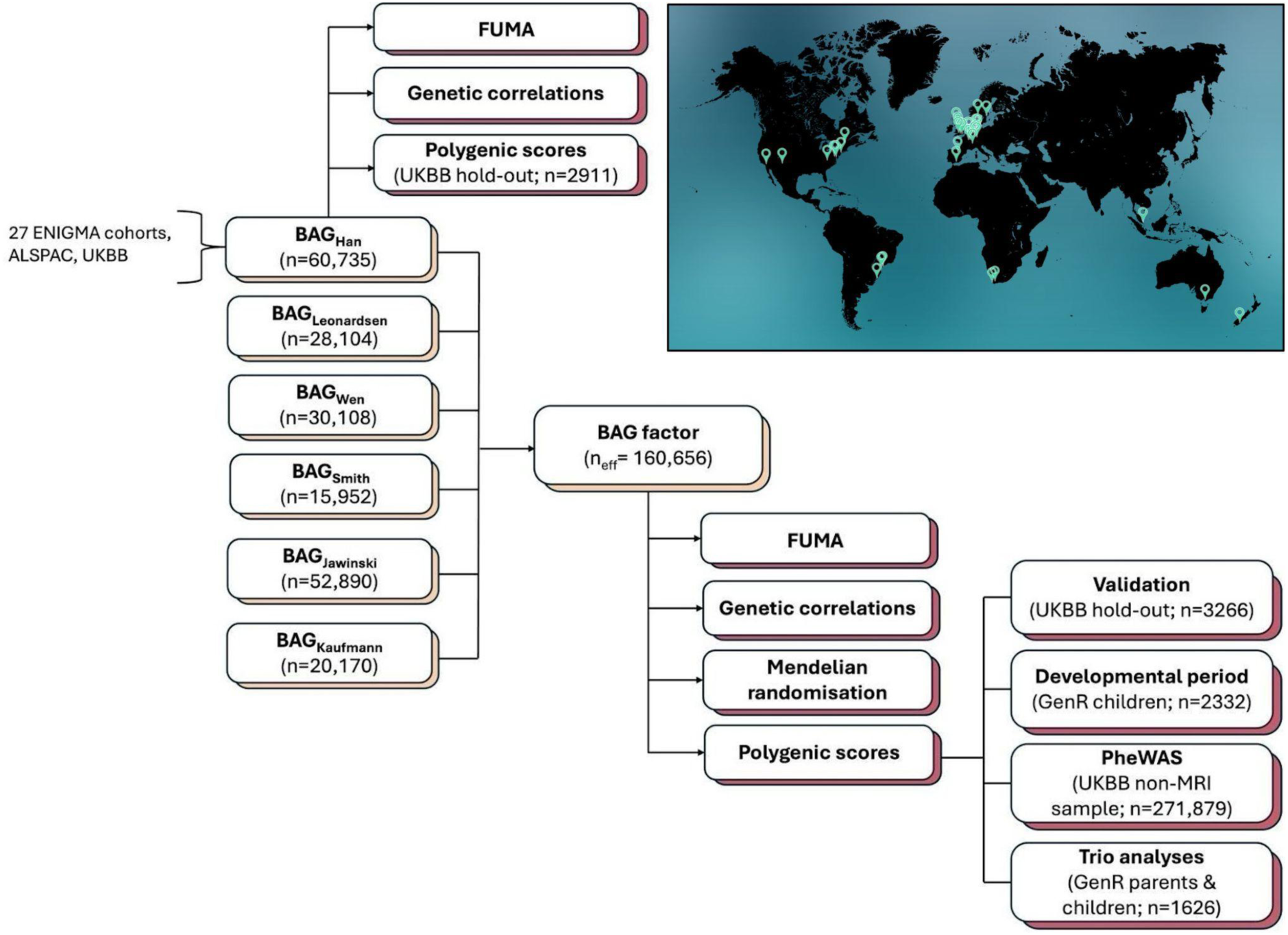
Analytical flowchart of the current study *Note*. Light brown boxes represent genome-wide association studies (GWASs). Maroon boxes represent post-GWAS analyses. Summary statistics for BAG_Han_ have been obtained as part of the present study (see map in the top right). Summary statistics for BAG_Leonardsen_, BAG_Wen,_ BAG_Smith_, BAG_Jawinski_, and BAG_Kaufmann_ have been obtained from previously published studies. BAG = brain age gap; PheWAS = phenome-wide association study; UKBB = UK Biobank; GenR = Generation R study.

### Genomic SEM GWAS of six brain age gaps

We used Genomic SEM (v.0.0.5; Grotzinger et al., 2019) to conduct a GWAS of the six BAGs listed in **Table 1** (*N*_unique.individuals_=62,568; *N*_effective_=160,656). This method accounts for sample overlap across contributing GWAS studies. First, we estimated genetic correlations between the six BAG models within genomic SEM and performed confirmatory factor analysis to identify a common latent factor. This approach allowed us to capture shared genetic variance while reducing measurement error and model-specific noise. We then regressed each SNP on the latent brain age factor using Genomic SEM’s multivariate GWAS function. For a detailed description of the method, see Grotzinger et al. (2019).

To evaluate the adequacy of a single-factor solution, we additionally tested an exploratory two-factor model and compared fit indices (standardised root mean square residual; SRMR, Comparative Fix Index; CFI, model χ² and Akaike Information Criterion; AIC) using confirmatory factor analysis. All modelling was conducted using default settings unless otherwise specified.

### Post-GWAS analyses

We carried out a comprehensive set of post-GWAS analyses, as detailed below. For more information on these post-GWAS analyses, see **SM section 1.2**.

#### Genomic loci and functional annotation

To annotate, prioritise and visualise GWAS results, we used the Functional Mapping and Annotation of Genome-Wide Association Studies (FUMA v1.5.2; Watanabe et al., 2017) *SNP2GENE* pipeline.

#### Gene functional annotation

FUMA’s *GENE2FUNC* pipeline provided biological context for the identified genes. Tissue-specific differentially expressed genes were identified by testing for enrichment of prioritised genes against background gene sets.

#### MAGMA gene-based, gene-set, and tissue expression analyses

Gene-based association analysis was performed using MAGMA v1.08. Gene-set analysis was performed for curated gene sets and GO terms obtained from the MsigDB database (Liberzon et al., 2011).

#### Heritability and genetic correlations

To perform heritability and genetic correlation analyses, we used linkage disequilibrium (LD) score regression (Bulik-Sullivan et al., 2015; Bulik-Sullivan et al., 2015). We assessed correlations of the brain age factor with 33 psychiatric, cardiometabolic, behavioural, and ageing-related traits.

#### Mendelian randomisation

To understand which traits may be causally related to BAG, we performed bi-directional two-sample Mendelian randomisation (MR) analysis with all 33 traits. An inverse-variance weighted (IVW) approach was used as our primary analysis estimate, with MR-Egger and weighted-median methods included as sensitivity analyses. To test the causal direction of each SNP and remove potentially invalid genetic variants, Steiger filtering was applied (Hemani et al., 2017). Analyses were performed using the TwoSampleMR package (Hemani et al., 2018) in R v4.3.2 (R Core Team, 2025). To account for sample overlap between exposure and outcome GWASs, we also calculated bias-corrected IVW estimates using the MRlap package (Mounier & Kutalik, 2023).

#### Polygenic score associations with BAG in childhood and adulthood

Polygenic scores (PGS) for individual BAGs and the latent BAG factor were computed using SBayesRC (Zheng et al., 2024). We assessed how well PGS scores predicted phenotypic BAGs **(1)** in a longitudinal child cohort (The Generation R Study; n = 2,332; mean age 10 years [SD = 0.61]; Kooijman et al., 2016) and **(2)** in an independent (hold-out) adult imaging subset from the UKBB cohort stratified by ancestry group (UKBB return no. 2442): European (n=2,000), Central/South Asian (n=638), East Asian (n=291), and African (n=337); mean age 65 years [range: 47-82]; **Table S1**). UKBB results for the polygenic BAG factor score were compared against scores obtained for each of the six input GWASs that contributed to the latent BAG factor. To ensure independence from the previous UKBB imaging GWASs, all participants who took part in the imaging visit and their relatives (up to 3rd degree) were excluded.

Due to data availability, we were only able to validate the PGS for the BAG factor using two phenotypic BAGs in the UKBB cohort (BAG_Han_ and BAG_Jawinksi_) and three in GenerationR (BAG_Han_, BAG_Leonardsen_, and one external model not included in the latent factor: BAG_Cole_; Cole, 2023).

#### Polygenic score associations with health traits in childhood and adulthood

We then tested for PGS associations with nine available health-related outcomes during development, using data from Generation R (Kooijman et al., 2016). Additionally, to test which BAG PGS (factor-based vs. individual PGSs) is most relevant to a wide range of traits (n_traits_=6,646), we performed a phenome-wide association study (PheWAS) in an independent, non-imaging subset of unrelated White British adults from the UKBB cohort (n=271,879; mean age 58 years [range: 40-74]). For PGS covariates see **SM Methods section 1.2**.

#### Trio genetic analyses

Within-family trio models were used to test for direct and indirect genetic effects on nine available health-related outcomes and three phenotypic BAGs in Generation R (Kooijman et al., 2016). In trio models, the direct genetic effect represented the effect of the child’s PGS on health outcomes (conditional on the parental PGSs), while the indirect effects reflected the parental effect on child health outcomes (conditional on the child’s PGS) (Pingault et al., 2023; Tubbs et al., 2020).

The analytical sample included genotyped trios (children and their biological mothers and fathers, up to n=1,063) with available phenotypic data. For each parent-offspring trio, we computed polygenic BAG factor scores using SBayesRC (Zheng et al., 2024). Before analysis, child, mother, and father polygenic BAG factor scores were first standardised and then residualised for the first five genetic principal components. The three polygenic BAG factor scores were used as predictors of three phenotypic BAGs in the offspring (BAG_Han_, BAG_Leonardsen_, and BAG_Cole_) as well as nine health outcomes in separate linear regression models. All outcomes were measured when the children were on average 9-13 years old. Each model also included age and sex as covariates.

## Results

### BAG_Han_ GWAS meta-analysis

We performed a genome-wide association meta-analysis of BAG_Han_ across 29 cohorts (total *N* = 60,735; mean age=37.6 years [range=18-75]; 53% female). We identified 32 lead SNPs across 29 genomic risk loci reaching genome-wide significance (*p* < 5e⁻⁸; **Table S5**). Mapped genes (**Table S6**-**S7**) have been previously associated with brain-related morphology (i.e., brain region volumes), cognitive and psychiatric traits (e.g., cognitive performance, schizophrenia), and other health-related measures (e.g., body mass index, lipids; **Table S11**). Polygenic scores based on BAG_Han_ explained the largest proportion of variance in brain age gap in European ancestry individuals (R^2^=8.3%; *n*=1,738) compared to other ancestries (R^2^<2.7%; **Table S28**). A full description of the BAG_Han_ GWAS results is provided in the **SM Results section 2.1** and **Tables S5–S12**.

### Shared genetic architecture across six brain age models

Genetically, all BAG models were significantly correlated with each other, with at least a moderate effect size (mean *r*_g_=0.64, **Figure 2A**; **Table S13**). Correlations ranged from *r*_g_=0.37 (BAG_Leonardsen_ and BAG_Han_) to *r*_g_=0.93 (BAG_Wen_ and BAG_Smith_). All six BAG models loaded onto a single latent factor (BAG_factor_; all standardised loadings > 0.60). This one-factor solution fitted the data well (χ2[9] = 77.14, *p* = 5.96^-13^, CFI = 0.94, SRMR = 0.09), explaining 63% of the total genetic variance. BAG_Wen_ had the highest factor loading (beta=0.91, SE=0.07, *p*=6.93e^-41^), followed by BAG_Smith_ and BAG_Kaufmann_ (**Figure 2B**). For the two-factor solution, see **SM Results section 2.2** and **Figure S3**.

**Figure 2.**
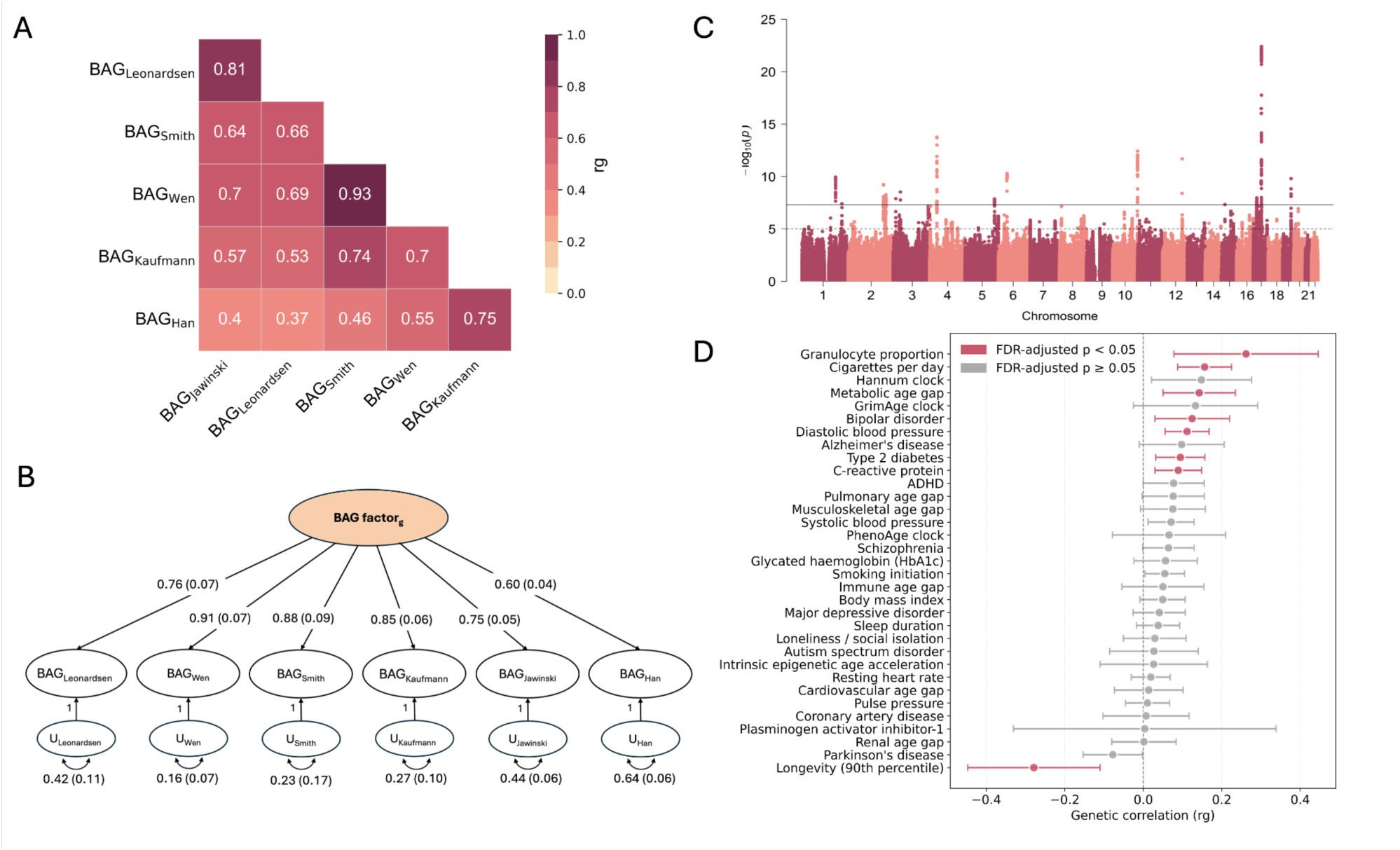
Shared genetic architecture of six brain age gaps and their association with health outcomes *Note*. (A) Genetic correlations among six brain age gaps. (B) Genetically defined common factor model for brain age gap with standardised coefficients and standard errors shown in brackets. The latent factor is specified with unit variance identification. Model fit metrics: χ² = 77.14, p(χ²) = 5.96e^-13^, CFI = 0.94, SRMR = 0.09. (C) Manhattan plot of the brain age gap GWAS based on genomic SEM using DWLS estimation. The y-axis depicts -log_10_(*p*) values for genetic variants associated with the brain age factor. The genome-wide significance threshold is denoted by the horizontal dotted red line at *p* = 5e^-8^. (D) Genetic correlations of the brain age factor with health outcomes. BAG = brain age gap; GWAS = genome-wide association study; SEM = structural equation model; DWLS = Diagonally Weighted Least Squares. ADHD = attention deficit hyperactivity disorder; PAI1 = DNA methylation-based estimator of plasminogen activator inhibitor-1.

### Genome-wide associations for shared genetic BAG factor

We identified 19 independent genome-wide significant SNPs (**Figure 2C**) across 16 independent loci (**Table 2**). The GWAS yielded a genomic control lambda (λGC) of 1.21 and an LD score regression intercept of 1.01 (SE=0.01), indicating inflation driven primarily by polygenicity.

**Table 2.**
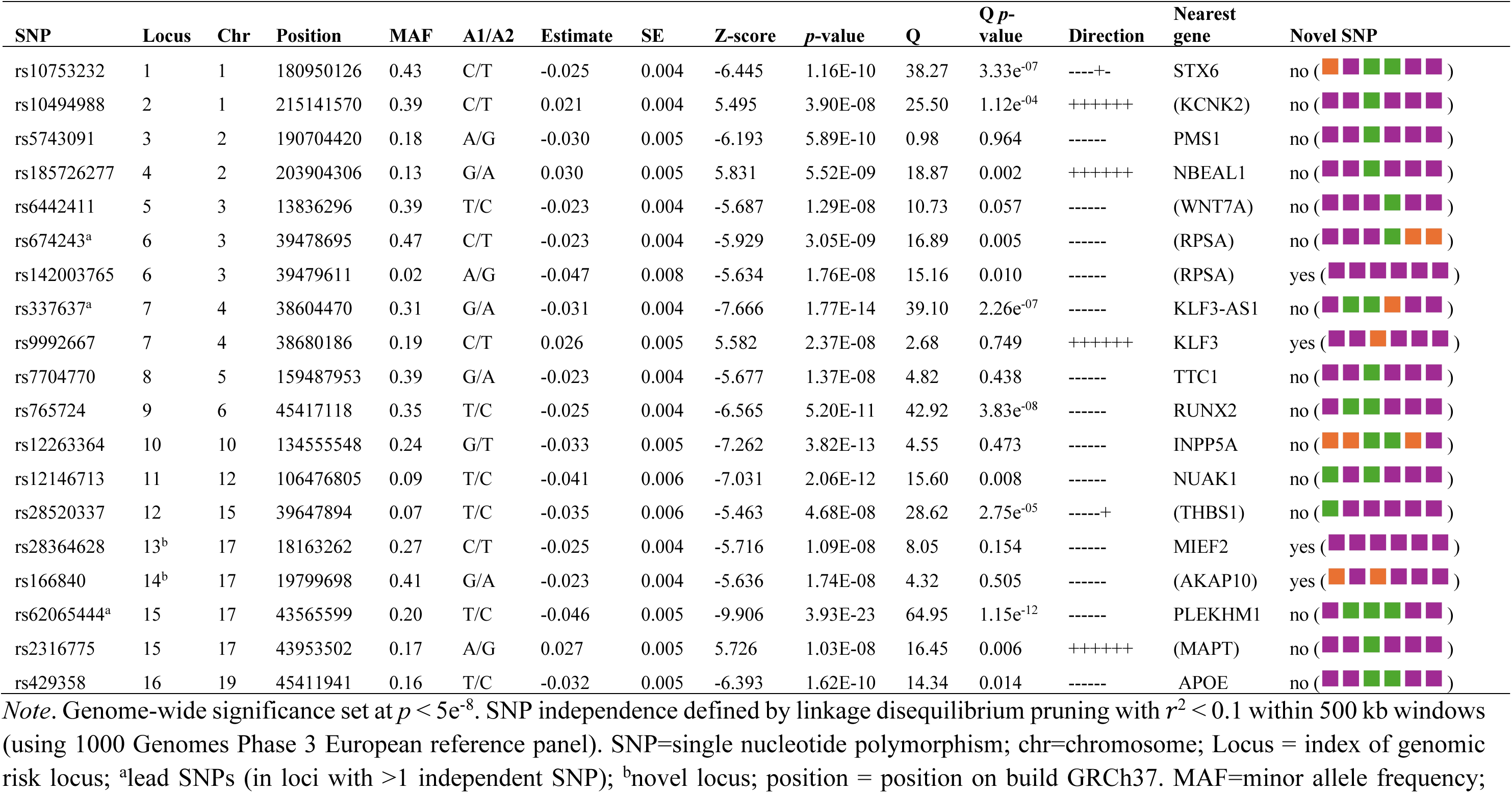
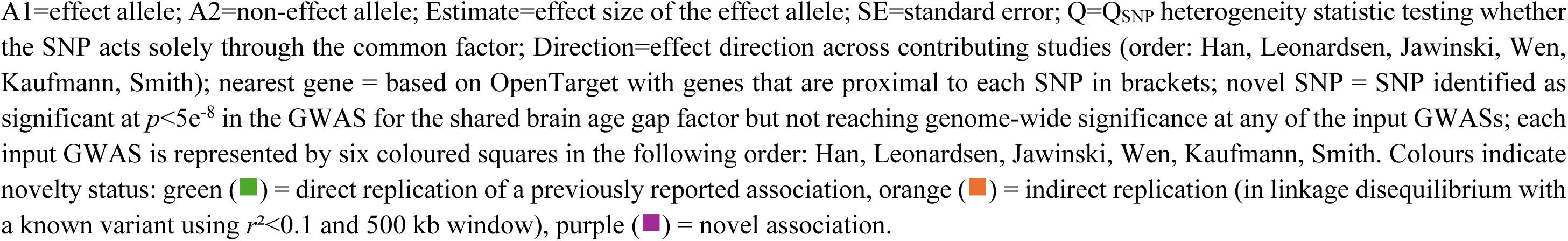
Nineteen independent genome-wide significant variants associated with brain age gap factor across 16 genomic loci.

Seventeen of the 19 independent SNPs showed a consistent direction of effect across contributing GWASs; 15 SNPs had been previously linked to BAG in the input GWASs, and four SNPs were novel (**Table 2**). The SNPs mapped most consistently to 32 genes, with *STH*, *MAPT,* and *NKX6-2* showing the strongest associations (all *p* < 9.69e^-13^; **Tables S14-S15**). A subset of these genes (e.g., *MAPT*, *KANSL1*, *CRHR1*, and *TOMM40*) had previously been linked with neurodegeneration, brain structure, or brain age (Burggren et al., 2017; Jawinski et al., 2025; Kulminski et al., 2025; Leonardsen et al., 2023; Zhang et al., 2016), whereas others (e.g., *SHMT1*, *ALKBH5*, *WDR12*) had no strong prior relation with brain age-related phenotypes, representing potentially novel genes underlying brain age.

### Gene functional annotation

We observed tissue-specific expression patterns, whereby some genes (e.g., *MAPT*, *RUNDC3A*, and *GFAP*) showed higher expression in brain tissue compared to peripheral tissues (**Figure S4**, **Table S16**). The strongest enrichment signals were observed in the cerebellum, not significant after Bonferroni correction (**Figure S5**, **Table S17**). Enriched biological processes were related to tumour immune responses (**Table S18**). Lastly, among all traits included in the GWAS catalog, the prioritised genes overlapped most significantly with phenotypes related to neurodegenerative disease, cardiometabolic health, and brain morphology (e.g., Alzheimer’s disease, body mass index, lipids, hippocampal volume, and cerebrospinal fluid biomarkers). Reassuringly, brain age was also identified among the enriched traits (**Figure S6**, **Table S18**-**S19**).

### Genetic correlations with other traits

The BAG factor was genetically correlated with 12 of 33 tested traits, of which eight survived false discovery rate (FDR) correction (**Figure 2D**; **Table S20**). These included positive associations with granulocyte proportion, smoking, metabolic age, bipolar disorder, blood pressure, type 2 diabetes, and CRP, as well as negative associations with longevity.

### Mendelian randomisation

MR results demonstrated evidence for a potentially causal role of blood pressure (systolic blood pressure: beta_IVW_ 0.066, 95% CI 0.017 to 0.116, *p*=0.008; diastolic blood pressure: beta_IVW_ 0.069, 95% CI 0.021 to 0.116, *p*=0.005), smoking initiation (beta_IVW_ 0.070, 95% CI 0.014 to 0.127, *p*=0.014), and longevity (OR_IVW_ 0.95, 95% CI 0.92 to 0.99, *p*=0.019) on the BAG factor (**Figure 3**; **Table S21**). Reverse MR revealed an effect of the BAG factor on autism spectrum disorder (OR_IVW_=1.85, 95% CI 1.29 to 2.66, *p* < 0.001), sleep duration (beta_IVW_=-0.094, 95% CI -0.163 to -0.026, *p* = 0.007), and smoking initiation (beta_IVW_=-0.144, 95% CI -0.248 to -0.041, *p* = 0.006). The MR-Egger estimate for smoking initiation was inconsistent with the primary estimate. No other trait showed significant associations (**Figure 3**; **Table S22**). For sensitivity analyses, see **SM Results section 2.3** and **Tables S21-S24**.

**Figure 3.**
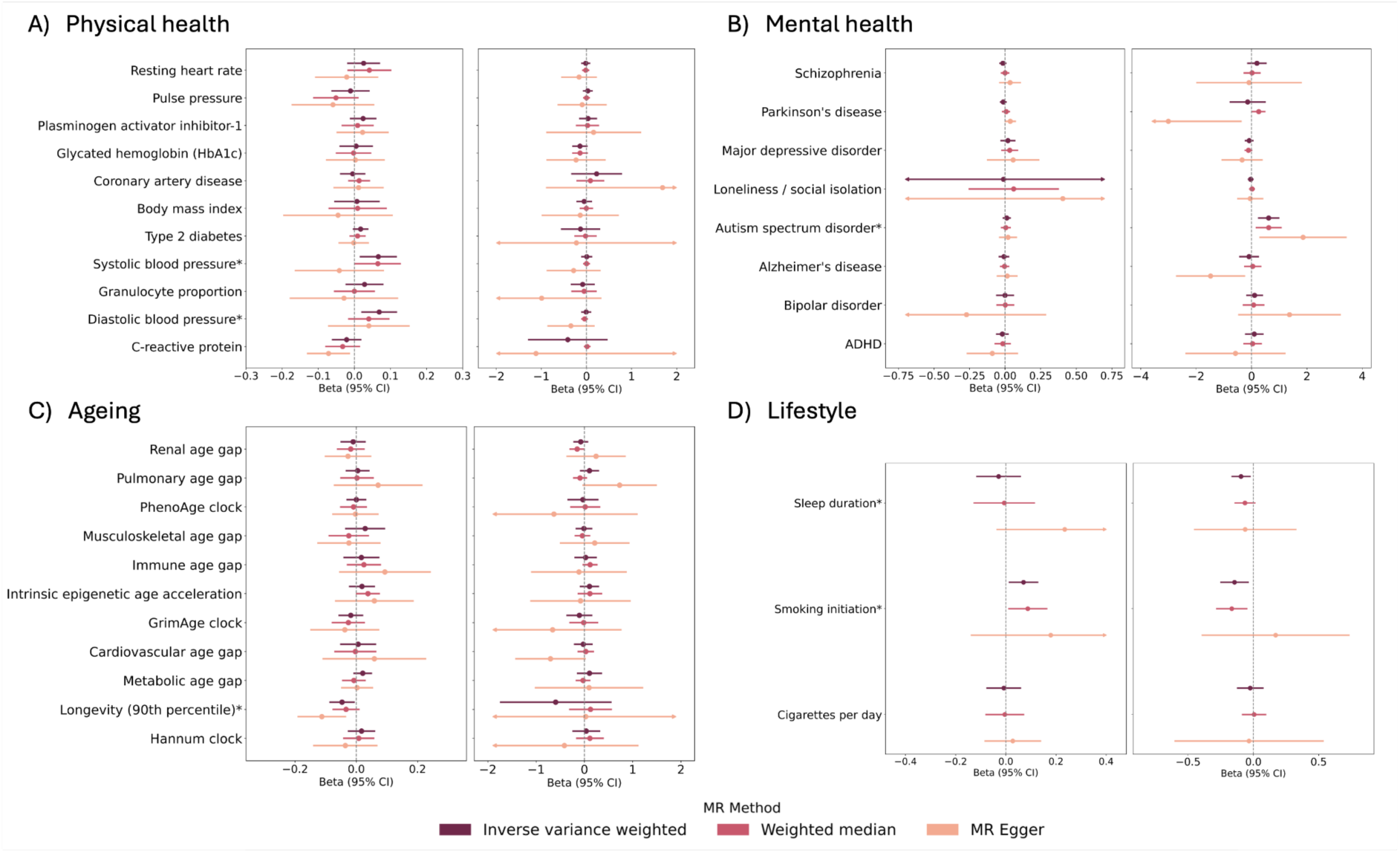
Mendelian randomisation results: bidirectional association between brain age gap factor and health outcomes *Note*. Each panel portrays the effect of health outcomes on brain age gap (left side) and the effect of brain age gap on health outcomes (right side). Beta (95% CI) represents the SD change in brain age gap per 1 SD increase in exposure. For binary outcomes (type 2 diabetes, coronary artery disease, schizophrenia, Parkinson’s disease, major depressive disorder, autism spectrum disorder, Alzheimer’s disease, bipolar disorder, ADHD, longevity, smoking initiation), effect estimates are presented on the log-odds scale. ADHD = attention deficit hyperactivity disorder; SD = standard deviation.

When compared to MR analyses conducted separately for each of the six individual BAGs, we observed that the BAG factor captured the most robust associations across traits (i.e., identified by most BAG models) and omitted inconsistent associations (i.e., only identified with one or two individual BAG models; **Figure S7**; **Table S21**). The same was largely true for the reverse associations (trait on BAG; **Figure S8**; **Table S22**). MR results for the BAG factor did not seem to be driven by biases related to sample overlap or winner’s curse. For details, see **SM Results section 2.3**, **Figure S9**; and **Table S24.**

### Polygenic score associations with BAG in childhood and adulthood

We then evaluated the performance of the polygenic score for the BAG factor in predicting phenotypic BAG in childhood and adulthood (including analyses across ancestries), using a sample of children (mean age 9-13 years; **Table S25**) from the Generation R Study and a multi-ancestry sample of adults from the UKBB cohort (mean age 65 years; **Table S26**). These samples were independent from the discovery GWAS.

In Generation R, small positive associations between the BAG factor polygenic score and phenotypic BAGs (BAG_Han_ and an external model BAG_Cole_) were observed, suggesting genetic influences on brain age already early in life (partial *R*^2^ up to 1.5%; **Figure 4A**; **Table S27**).

**Figure 4.**
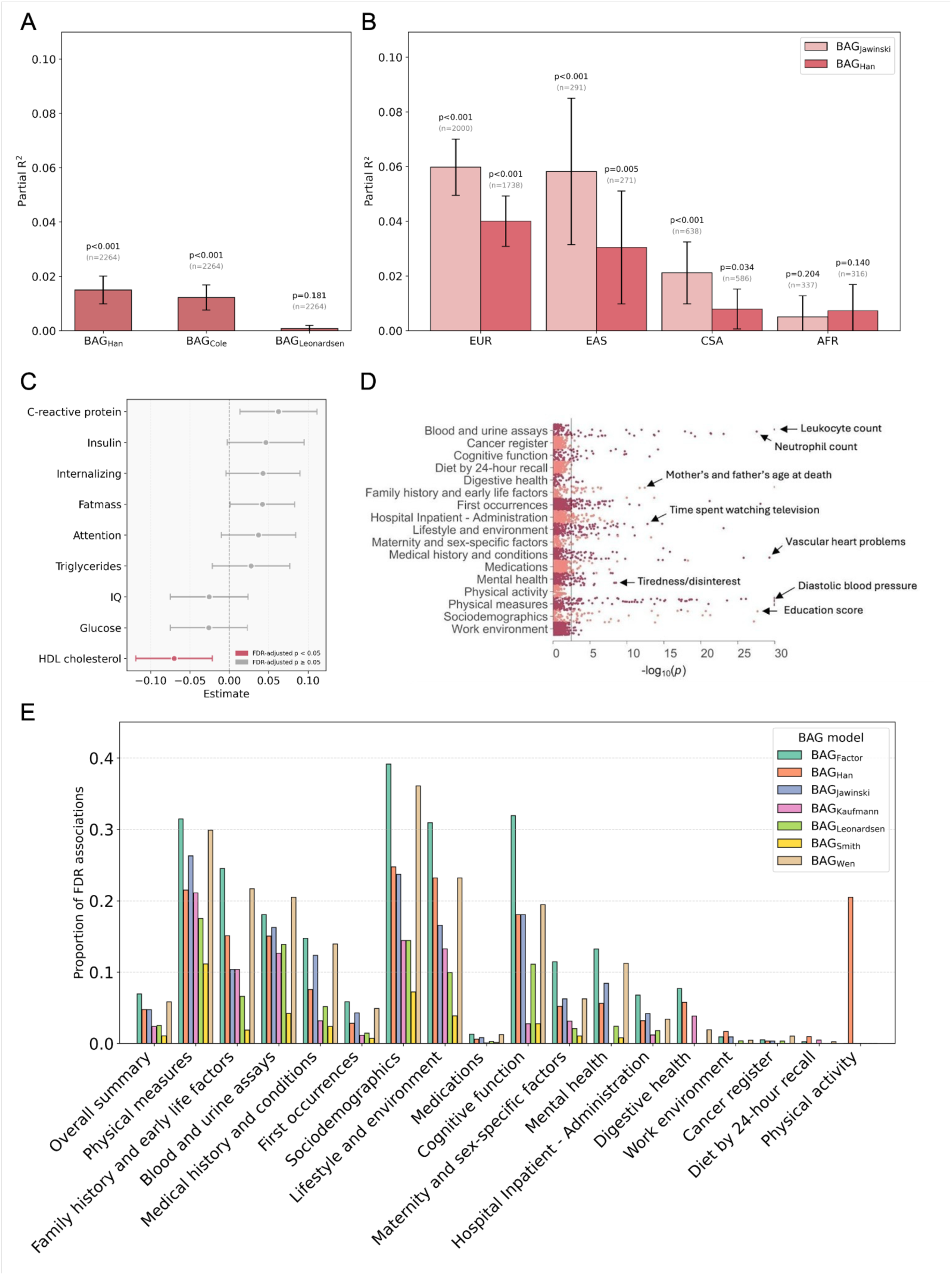
Associations of the polygenic score for BAG factor with phenotypic BAGs and a broad range of outcomes in the Generation R and UK Biobank cohorts *Note*. Association of the polygenic score for the BAG factor with **(A)** three phenotypic BAGs in children from the Generation R cohort, and **(B)** two phenotypic BAGs in adults from the UK Biobank cohort stratified by ancestry: European (EUR), East Asian (EAS), Central/South Asian (CSA), and African (AFR). Error bars represent standard errors. (**C**) Associations of the polygenic score for the BAG factor with nine health outcomes in Generation R. **(D)** PheWAS results showing associations of the polygenic score for BAG factor with a broad range of outcomes in the UK Biobank cohort, with the top (or most relevant) association in each category annotated by an arrow. **(E)** Proportion of PheWAS associations surviving FDR correction across phenotypic categories for each polygenic score derived from BAG_factor_ and the six input GWASs that contributed to the meta-analysis. BAG = brain age gap; PheWAS = phenome-wide association study; GWAS = genome-wide association study; FDR = False Discovery Rate.

In UKBB, positive associations were observed between the polygenic score for the BAG factor with both BAG phenotypes (i.e., BAG_Han_ and BAG_Jawinski_), showing the best predictive performance in European and East Asian ancestry groups (partial *R*^2^ up to 6%; **Figure 4B**). Cross-model analyses showed that the best-matching polygenic score-BAG pairs used the same model (i.e., PGS_BAG.Han_ with BAG_Han_ and PGS_BAG.Jawinski_ with BAG_Jawinski_). When removing the exact pairs, the BAG factor consistently ranked highest, explaining the largest proportion of variance across the remaining BAG phenotypes and ancestry groups (**Table S28**).

### Polygenic score associations with health traits in childhood and adulthood

When testing for associations between the PGS for the BAG factor and nine health-related child outcomes in Generation R (n=2,269), we observed a negative association with high-density lipoprotein cholesterol, which survived multiple testing correction (**Figure 4C, Table S29**). No other trait showed significant associations. In UKBB (n=271,879), the PGS for the BAG factor was associated with 460 adult phenotypes in a PheWAS after FDR correction. Notable associations included blood pressure, education/qualification, overall health rating, parental age at death, fluid intelligence score, neuroticism, and alcohol use (**Figure 4D**). Overall, the PGS for the BAG factor outperformed PGSs for the individual BAG models that contributed to the meta-analysis, achieving the lowest mean rank of association *p*-values across phenotypes and the highest number of significant associations compared to polygenic scores from individual BAG models (**Figure 4E**; **Figure S10**; **Table S30**). Across all BAG models, the highest proportions of FDR-corrected associations were in domains related to sociodemographic, lifestyle/environment, physical, and cognitive characteristics. BAG_factor_ showed the highest number of associations within most categories, indicating the broadest phenotypic coverage amongst all BAGs. A detailed breakdown by phenotype category (e.g., mental health, cognitive function, medications, physical measures, sociodemographic characteristics) is provided in **Table S31**.

### Trio genetic analyses

Finally, we investigated whether associations between the PGS for the BAG factor and health-related child outcomes in Generation R were likely due to indirect genetic effects, using child-parent trio data (up to n=1,063). We detected no significant indirect genetic effects for any trait (**Figure 5; Table S32**).

**Figure 5.**
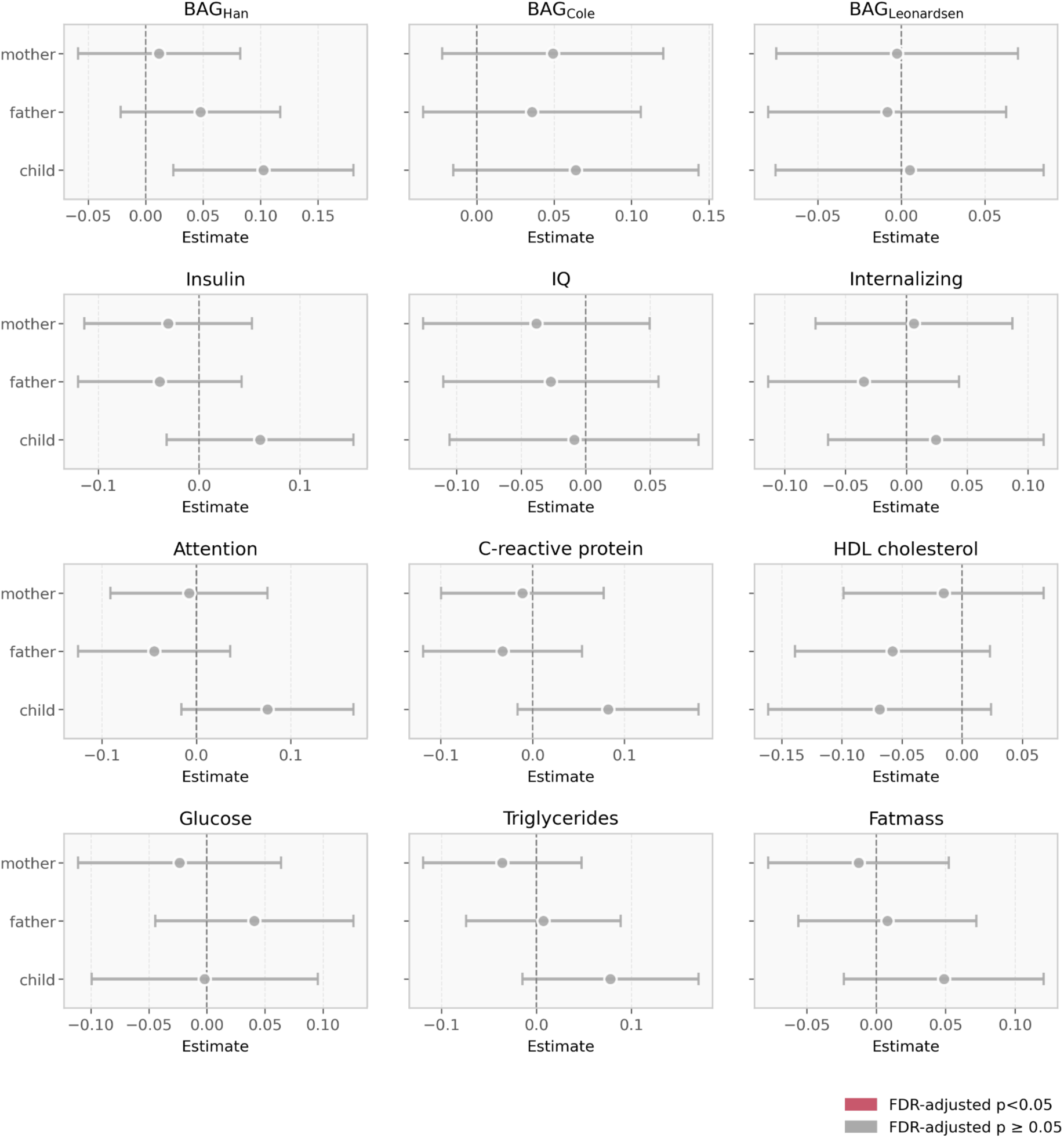
Trio results testing for indirect and direct (child) genetic effects of polygenic score for BAG factor on child outcomes in Generation R *Note*. IQ = intelligence quotient; BAG = phenotypic brain age gap.

## Discussion

Our genomic analysis demonstrated that diverse BAG models, trained on distinct imaging modalities, features, and samples, were largely underpinned by a common genetic signal. All six BAG GWASs loaded strongly on a single latent factor that accounted for ∼63% of their additive genetic variance (range_individual.BAGs_: 36-83%), indicating that a sizable proportion of genetic influences on brain age is shared across methodological implementations. A GWAS of this factor yielded 19 independent variants, of which four were novel and two were independent of known loci. The remaining 15 variants had been previously identified in one to three BAG GWASs, yet never consistently across all six GWASs. These variants map to genes previously implicated in neurodegeneration and brain structure (e.g., *MAPT*, *KANSL1*, *CRHR1*, *APOE*/*TOMM40*). In line with previous studies (Jawinski et al., 2025; Leonardsen et al., 2023), the strongest signal was observed at a locus linked to *MAPT* – a gene encoding tau, a key protein in Alzheimer’s disease and other tauopathies (Zhang et al., 2024). We also observed signals in novel candidates (e.g., *SHMT1*, *ALKBH5*, *WDR12*), which are involved in the regulation of cell proliferation (Hölzel et al., 2005) and apoptosis (Paone et al., 2014). These results point to a biologically plausible, polygenic substrate of brain age that is shared but only partially recoverable from any single BAG implementation.

Further analyses position the latent BAG factor as a health-relevant dimension. Genetic correlations aligned BAG with cardiometabolic and inflammatory profiles (blood pressure, type 2 diabetes, C-reactive protein, granulocyte proportion), smoking, bipolar disorder, and metabolic age, and in the expected opposite direction with longevity. MR supported putative causal contributions of higher blood pressure and smoking initiation to higher BAG (pointing towards promising prevention targets for brain health), and of a genetic propensity to longevity as a protective factor. The implication of blood pressure on brain age replicates previous MR findings (Jawinski et al., 2025). Smoking initiation and longevity represented associations not reported in previous MR studies, but are in line with those observed in non-causal observational research (Linli et al., 2022; Zettergren et al., 2024). Reverse MR indicated a possible causal effect of genetic liability to higher BAG factor on autism spectrum disorder and shorter sleep duration, suggesting that BAG may be both a downstream effect of and a risk factor for poor health.

Polygenic scores derived from the latent factor predicted BAGs in independent cohorts, including in childhood, suggesting that brain age trajectories may be shaped well before midlife. Most ageing studies concentrate on adulthood (Felix et al., 2014), despite recent research showing that variations in adult brain age might be shaped by early-life experiences (Vidal-Pineiro et al., 2021). Consistent with this research, our findings call for an increased focus on development to understand lifelong ageing and to identify effective ageing interventions.

In our PheWAS analyses, polygenic scores derived from the latent factor also outperformed scores from individual BAG GWASs in predicting health traits across multiple domains. This increase in phenotypic associations when combining different GWASs has also been observed in the context of substance use (Xu et al., 2023) and cardiovascular disease (Jorda et al., 2023; Jordà et al., 2025), which further highlights the interconnectedness of a wide range of phenotypes and genetically indexed brain age. These findings suggest that BAG may reflect broad systemic effects and therefore serve as a more global marker of health, rather than being limited to brain-related traits (Cole et al., 2018).

Trio models suggested that associations of the polygenic score for the BAG factor with the individual BAGs and HDL cholesterol in childhood were unlikely to reflect intergenerational pathways. The overall lack of effects aligns with prior research (Ghatan et al., 2025), although due to the small sample size, we might have been underpowered to detect these effects, or they may emerge later.

Our findings should be interpreted in light of the following limitations. First, while the latent genetic BAG factor removes model-specific measurement error, it may still partly capture shared methodological characteristics rather than solely biological brain age processes. Second, due to the limited availability of MRI data in non-European cohorts, our analyses were based on predominantly European-ancestry individuals, although we did include cross-ancestry analyses where possible. Third, our BAG factor was largely T1-based, potentially underestimating vascular effects on the brain that are better captured by T2 or FLAIR sequences. Future studies could build on our study by assessing harmonised, region- or modality-specific brain age models (e.g., structural vs. functional, or grey vs. white matter) and examining effects across multiple ancestries. Findings could also be triangulated across longitudinal designs and in randomised controlled trials to test whether blood pressure treatment or smoking cessation could slow down BAG trajectories.

In conclusion, by modelling shared genetic variance across six distinct brain age models, this study provides a comprehensive and biologically grounded understanding of the genetic architecture underlying brain ageing. The identification of 19 genome-wide significant SNPs––spanning established neurodegenerative genes (e.g., *MAPT*, *APOE/TOMM40*) and novel candidates––highlights a core set of genetic influences that transcend methodological differences between BAG implementations. The latent BAG factor not only enhanced genetic discovery and predictive accuracy but also captured meaningful associations with health-relevant traits, including blood pressure, smoking, and longevity, underscoring its systemic relevance. Polygenic analyses further revealed that genetic influences on brain age gap may emerge early in life. Collectively, these findings position the latent BAG factor as a robust, integrative phenotype that advances our understanding of the biological foundations of brain age gap and its links to broader health outcomes.

## Code availability

All analysis code is available at https://github.com/VilteBaltra/genetic-architecture-of-brain-age-gap, with UK Biobank–specific code for brain age estimation, GWAS, and PheWAS provided at https://github.com/pjawinski/enigma_brainage.

## Data Availability

GWAS summary statistics for BAG_Leonardsen_ are available from the corresponding author of the associated publication (Yunpeng Wang; doi: 10.1038/s41380-023-02087-y) upon reasonable request. GWAS summary statistics for BAG_Wen_ can be downloaded from https://labs-laboratory.com/medicine/multimodal_brain_bag. GWAS summary statistics for BAG_Smith_ can be obtained from https://www.fmrib.ox.ac.uk/ukbiobank/BrainAgingModes (model number: V0140). GWAS summary statistics for BAG_Kaufmann_ can be obtained from https://github.com/tobias-kaufmann/brainage. GWAS summary statistics for BAG_Jawinski_ can be downloaded from Zenodo at https://doi.org/10.5281/zenodo.14826943. We used the *brainage2025.full.eur.gwm.gz* dataset comprising the discovery and replication sample (n = 54,890; European ancestry), excluding 2,000 individuals that were held out for polygenic score analyses, resulting in 52,890 individuals of European ancestry. GWAS summary statistics for the ENIGMA brain age model (BAG_Han_) and the latent brain age gap factor (BAG factor) have been deposited in Zenodo and are available at https://doi.org/10.5281/zenodo.17997141 and https://doi.org/10.5281/zenodo.17992281, respectively. Access is currently restricted while the associated manuscripts are under peer review and will be made publicly available upon publication.

## Acknowledgments

For cohort-specific funding and acknowledgements, see **SM Table S1**. MAN received funding through a Juan de la Cierva Contract (FJC2021-047538-I), funded by the Agencia Estatal de Investigación (AEI), Ministerio de Ciencia, Innovación y Universidades, Spain. OA was supported by the Research Council of Norway/Norges forskningsråd (Grant numbers: 324499, 324252, 296030); Nordforsk: (Grant number: 164218); South-East Norway Health Authority/Helse Sør Øst (Grant number: 2023-031). RAB was supported by the São Paulo Research Foundation (FAPESP), National Research Council CNPq. EMB was supported by the São Paulo Research Foundation (FAPESP; grant #2023/16061-0). CC was supported by EU HorizonEurope (FAMILY, grant agreement No 101057529). DD is partially supported by National Institute for Health and Care Research (NIHR) Maudsley Biomedical Research Centre, South London and Maudsley NHS Trust. The views expressed are those of the author(s) and not necessarily those of the NIHR or the Department of Health and Social Care. JF was supported by 1IK6CX002519 Research Career Scientist, Veterans Administration. AF was supported by NIH grant [R01AG073593]. DG was supported by National Institute of Mental Health grants MH078143, MH078111, and MH083824. LH was funded by the Rubicon (grant number 452020227) and Veni award (grant number 09150162210201) from the Dutch Research Council (NWO). LTI is supported by the São Paulo Research Foundation (FAPESP; grant #2022/15880-5). This work was supported by the National Center for Research Resources at the National Institutes of Health [grant numbers: NIH 1 U24 RR021992 (Function Biomedical Informatics Research Network), NIH 1 U24 RR025736-01 (Biomedical Informatics Research Network Coordinating Center; http://www.birncommunity.org], and the National Institute of Mental Health [grant number: R01 MH094524]. TGMvE, AL, and SOMVK were supported by the National Institute of Mental Health of the National Institutes of Health under award number R01 MH1345261. DK was supported by Stress in Action – Innovation in the understanding and application of daily-life stress. which is financially supported by the Dutch Research Council and the Dutch Ministry of Education, Culture and Science (NWO gravitation Grant No. 024.005.010). CL was supported by the Swedish Research Council (2022-01188), Swedish Brain Foundation (FO2025-0276-HK-280). ILC was supported by Fundacao de Amparo a Pesquisa do Estado de Sao Paulo (FAPESP), grant number: 2013/07559-3, Conselho Nacional de Pesquisa (CNPq), Brazil (grant number 311923/2019-4) and Coordenação de Aperfeiçoamento de Pessoal de Nível Superior (CAPES), Brazil (grant number 001). YM was partially supported by Amsterdam UMC (Starter Grant Ronde 2), Amsterdam Neuroscience (PoC funding 2024-2026), and the ImmunoMIND consortium, funded by UK Research and Innovation as part of the UK national Mental Health Platform. TP was supported by CIHR. RP has been funded by the New Zealand Health Research Council (program grant no. 16-604) and the New Zealand Ministry of Business, Innovation and Employment. AR was supported by National Institute of Mental Health grants MH078143, MH078111, and MH083824. DJS was funded by the Medical Research Council of South Africa. AT has been funded by the Deutsche Forschungsgemeinschaft (DFG, German Research Foundation) – 542489987. RT has been funded by the New Zealand Health Research Council (program grant no. 16-604) and the New Zealand Ministry of Business, Innovation and Employment. SHIP is part of the Community Medicine Research Network of the University Medicine Greifswald, which is supported by the German Federal State of Mecklenburg-West Pomerania. EW received funding from UK Research and Innovation (UKRI) under the UK government’s Horizon Europe / ERC Frontier Research Guarantee [BrainHealth, grant number EP/Y015037/1] and from Wellcome (reference 315898/Z/24/Z). JMW is part-funded by the UK Dementia Research Institute (funded by UK Medical Research Council, Alzheimer’s Society and Alzheimer’s Research UK), the National Institute of Health and Care Research; the work was part funded by the UK Medical Research Council, Biotechnology and Biological Sciences Research Council, AgeUK, and Row Fogo Charitable Trust. HJZ is funded by the Medical Research Council of South Africa. UKBB work was done under application no. 423032. LTW was supported by the Research Council of Norway (249795, 273345, 298646, 300768, 324499), the South-Eastern Norway Regional Health Authority (2018076, 2019101), and the European Research Council under the European Union’s Horizon 2020 Research and Innovation Programme (802998). SRC is supported by a Sir Henry Dale Fellowship jointly funded by the Wellcome Trust and the Royal Society (221890/Z/20/Z). AF is supported by NIH grant [1RF1AG073593]. Ms Kayleigh Beukes is acknowledged for assistance with data curation of the COMPIMP dataset.

## Conflict of interest

OAA has received speaker fees from BMS, Lundbeck, Janssen, Otsuka, Lilly, and Sunovion and is a consultant to Cortechs.ai. and Precision Health. JC is shareholder in and scientific advisor to BrainKey and Claritas HealthTech. YM received consulting fees to his institution from Noema Pharma outside the submitted work.

## Contribution Statement

Conceptualization: EW, DD, VB, JC, SEM.

Methodology: PJ, LH, LS, EW, DD, VB, MS, JBP, JC, CAMC, SEM.

Data analysis (individual cohorts): PK,BC,TF,MEB,MEB,FC,KS,RR,GAS,FC,YQ,CLY,MJC,HL,IL,DCG,TGMvE,AH,AK,AC, TM,KS,HJZ,YQ,GAS,OG,ARK,RAB,LKMH,JMW,BZK,AEF,SM,AL,EMB,SK,JS,DK,AJ, MS,CC,AT,SD,EW,SEM,AK,AT,MSM,KW,NP,YL,ÍK,GD,MA,IK.S,AM,MLS,LTI,JCI,OJ W, NWP, PJ.

Data curation and administration (individual cohorts): LTW,BWJHP,UV,RT,RP,MJG,KNTM,AC,OAA,TP,SRC,SB,YM,AT,SD,EW,SEM,MEB,M EB,FC,LS,KS,HJG,PMP,ZP,LH,RLM,MA,IKS,AM,MLS,LTI,JCI,OJW,RR,GAS,FC,YQ,C LY,MJC,HL,IL,DCG,TGMvE,AH,AK,AC,TM,KS,HJZ,YQ,GAS,OG,ARK,RAB,LKMH,JM W,ALR,DA,DJK,CAMC,SEM.

Quality control (individual cohorts): HV,LS,KS,HJG,PMP,ZP,LH,RLM,AK,AT,MSM,KW,NP,YL,ÍK,GD,MA,IKS,AM,MLS,LT I,JCI,OJW,PJ,KS,RR,GAS,FC,YQ,CLY,MJC,HL,IL,DCG,TGMvE,AH,AK,AC,TM,KS,HJZ,YQ,GAS,OG,ARK,RAB,LKMH,JMW, MS, YM, NWM.

Data acquisition (individual cohorts): DJS,CL,RS,SK,VC,DHM,JMF,RB,TEM,CL,PK,BC,TF,MEB,MEB,FC,KS,RR,GAS,FC,YQ, CLY,MJC,HL,IL,DCG,TGMvE,AH,AK,AC,TM,KS,HJZ,YQ,GAS,OG,ARK,RAB,LKMH,J MW,LTW,BWJHP,UV,RT,RP,MJG,KNTM,AC,OAA,TP,SRC,SB,HV,LS,KS,HJG,PMP,ZP,LH,RLM,HT.

Writing - original draft: VB, EW, DD.

Writing - Review & Editing: all authors.

Visualization: VB, SD, EW, PJ, ST.

## Notes

### Funding Statement

This study was funded through UK Research and Innovation (UKRI) under the UK government's Horizon Europe / ERC Frontier Research Guarantee [BrainHealth, grant number EP/Y015037/1]. For cohort-specific funding and acknowledgements, see SM Table S1. MAN received funding through a Juan de la Cierva Contract (FJC2021-047538-I), funded by the Agencia Estatal de Investigacion (AEI), Ministerio de Ciencia, Innovacion y Universidades, Spain. OA was supported by the Research Council of Norway/Norges forskningsrad (Grant numbers: 324499, 324252, 296030); Nordforsk: (Grant number: 164218); South-East Norway Health Authority/Helse Sor Ost (Grant number: 2023-031). RAB was supported by the Sao Paulo Research Foundation (FAPESP), National Research Council CNPq. EMB was supported by the Sao Paulo Research Foundation (FAPESP; grant #2023/16061-0). CC was supported by EU HorizonEurope (FAMILY, grant agreement No 101057529). DD is partially supported by National Institute for Health and Care Research (NIHR) Maudsley Biomedical Research Centre, South London and Maudsley NHS Trust. The views expressed are those of the author(s) and not necessarily those of the NIHR or the Department of Health and Social Care. JF was supported by 1IK6CX002519 Research Career Scientist, Veterans Administration . AF was supported by NIH grant [R01AG073593]. DG was supported by National Institute of Mental Health grants MH078143, MH078111, and MH083824. LH was funded by the Rubicon (grant number 452020227) and Veni award (grant number 09150162210201) from the Dutch Research Council (NWO). LTI is supported by the Sao Paulo Research Foundation (FAPESP; grant #2022/15880-5). This work was supported by the National Center for Research Resources at the National Institutes of Health [grant numbers: NIH 1 U24 RR021992 (Function Biomedical Informatics Research Network), NIH 1 U24 RR025736-01 (Biomedical Informatics Research Network Coordinating Center; http://www.birncommunity.org], and the National Institute of Mental Health [grant number: R01 MH094524]. TGMvE, AL, and SOMVK were supported by the National Institute of Mental Health of the National Institutes of Health under award number R01 MH1345261. DK was supported by Stress in Action - Innovation in the understanding and application of daily-life stress. which is financially supported by the Dutch Research Council and the Dutch Ministry of Education, Culture and Science (NWO gravitation Grant No. 024.005.010). CL was supported by the Swedish Research Council (2022-01188), Swedish Brain Foundation (FO2025-0276-HK-280). ILC was supported by Fundacao de Amparo a Pesquisa do Estado de Sao Paulo (FAPESP), grant number: 2013/07559-3, Conselho Nacional de Pesquisa (CNPq), Brazil (grant number 311923/2019-4) and Coordenacao de Aperfeicoamento de Pessoal de Nivel Superior (CAPES), Brazil (grant number 001). YM was partially supported by Amsterdam UMC (Starter Grant Ronde 2), Amsterdam Neuroscience (PoC funding 2024-2026), and the ImmunoMIND consortium, funded by UK Research and Innovation as part of the UK national Mental Health Platform. TP was supported by CIHR. RP has been funded by the New Zealand Health Research Council (program grant no. 16-604) and the New Zealand Ministry of Business, Innovation and Employment. AR was supported by National Institute of Mental Health grants MH078143, MH078111, and MH083824. DJS was funded by the Medical Research Council of South Africa. AT has been funded by the Deutsche Forschungsgemeinschaft (DFG, German Research Foundation) - 542489987. RT has been funded by the New Zealand Health Research Council (program grant no. 16-604) and the New Zealand Ministry of Business, Innovation and Employment. SHIP is part of the Community Medicine Research Network of the University Medicine Greifswald, which is supported by the German Federal State of Mecklenburg- West Pomerania. EW received funding from UK Research and Innovation (UKRI) under the UK government's Horizon Europe / ERC Frontier Research Guarantee [BrainHealth, grant number EP/Y015037/1] and from Wellcome (reference 315898/Z/24/Z). JMW is part-funded by the UK Dementia Research Institute (funded by UK Medical Research Council, Alzheimer's Society and Alzheimer's Research UK), the National Institute of Health and Care Research; the work was part funded by the UK Medical Research Council, Biotechnology and Biological Sciences Research Council, AgeUK, and Row Fogo Charitable Trust. HJZ is funded by the Medical Research Council of South Africa. UKBB work was done under application no. 423032. LTW was supported by the Research Council of Norway (249795, 273345, 298646, 300768, 324499), the South-Eastern Norway Regional Health Authority (2018076, 2019101), and the European Research Council under the European Union's Horizon 2020 Research and Innovation Programme (802998). SRC is supported by a Sir Henry Dale Fellowship jointly funded by the Wellcome Trust and the Royal Society (221890/Z/20/Z). AF is supported by NIH grant [1RF1AG073593]. Ms Kayleigh Beukes is acknowledged for assistance with data curation of the COMPIMP dataset.

### Author Declarations

All studies were conducted with appropriate ethical approval at each contributing site, and all participants provided written informed consent. UK Biobank received ethical approval from the National Research Ethics Service Committee North West-Haydock (reference 11/NW/0382) and this work was done under application no. 423032. Ethical approval for ALSPAC was obtained from the ALSPAC Ethics and Law Committee and Local Research Ethics Committees (e.g., Bristol and Weston Health Authority: E1808).

## References

Bacas, E., Kahhalé, I., Raamana, P. R., Pablo, J. B., Anand, A. S., & Hanson, J. L. (2023). Probing multiple algorithms to calculate brain age: Examining reliability, relations with demographics, and predictive power. Human Brain Mapping, 44(9), 3481–3492. 10.1002/hbm.26292

Baselmans, B. M. L., Yengo, L., Van Rheenen, W., & Wray, N. R. (2021). Risk in Relatives, Heritability, SNP-Based Heritability, and Genetic Correlations in Psychiatric Disorders: A Review. Biological Psychiatry, 89(1), 11–19. 10.1016/j.biopsych.2020.05.034

Bashyam, V. M., Erus, G., Doshi, J., Habes, M., Nasrallah, I. M., Truelove-Hill, M., Srinivasan, D., Mamourian, L., Pomponio, R., Fan, Y., Launer, L. J., Masters, C. L., Maruff, P., Zhuo, C., Völzke, H., Johnson, S. C., Fripp, J., Koutsouleris, N., Satterthwaite, T. D., … Davatzikos, C. (2020). MRI signatures of brain age and disease over the lifespan based on a deep brain network and 14 468 individuals worldwide. Brain, 143(7), 2312–2324. 10.1093/brain/awaa160

Blake, K. V. (2021). The brain age gap in social anxiety disorder. OpenUCT. http://hdl.handle.net/11427/35635

Bulik-Sullivan, B., Finucane, H. K., Anttila, V., Gusev, A., Day, F. R., Loh, P.-R., Duncan, L., Perry, J. R. B., Patterson, N., Robinson, E. B., Daly, M. J., Price, A. L., & Neale, B. M. (2015). An atlas of genetic correlations across human diseases and traits. Nature Genetics, 47(11), Article 11. 10.1038/ng.3406

Bulik-Sullivan, B. K., Loh, P.-R., Finucane, H. K., Ripke, S., Yang, J., Patterson, N., Daly, M. J., Price, A. L., & Neale, B. M. (2015). LD Score regression distinguishes confounding from polygenicity in genome-wide association studies. Nature Genetics, 47(3), 291–295. 10.1038/ng.3211

Cole, J. (2023). PyBrainAge. GitHub. https://github.com/james-cole/PyBrainAge

Cole, J. H., Ritchie, S. J., Bastin, M. E., Valdés Hernández, M. C., Muñoz Maniega, S., Royle, N., Corley, J., Pattie, A., Harris, S. E., Zhang, Q., Wray, N. R., Redmond, P., Marioni, R. E., Starr, J. M., Cox, S. R., Wardlaw, J. M., Sharp, D. J., & Deary, I. J. (2018). Brain age predicts mortality. Molecular Psychiatry, 23(5), 1385–1392. 10.1038/mp.2017.62

Desikan, R. S., Ségonne, F., Fischl, B., Quinn, B. T., Dickerson, B. C., Blacker, D., Buckner, R. L., Dale, A. M., Maguire, R. P., Hyman, B. T., Albert, M. S., & Killiany, R. J. (2006). An automated labeling system for subdividing the human cerebral cortex on MRI scans into gyral based regions of interest. NeuroImage, 31(3), 968–980. 10.1016/j.neuroimage.2006.01.021

Felix, J. F., Voortman, T., Van Den Hooven, E. H., Sajjad, A., Leermakers, E. T. M., Tharner, A., Jong, J. C. K., Duijts, L., Verhulst, F. C., De Jongste, J. C., Tiemeier, H., Hofman, A., Rivadeneira, F., Moll, H. A., Raat, H., Jaddoe, V. W., & Franco, O. H. (2014). Health in children: A conceptual framework for use in healthy ageing research. Maturitas, 77(1), 47–51. 10.1016/j.maturitas.2013.09.011

Fischl, B. (2012). FreeSurfer. NeuroImage, 62(2), 774–781. 10.1016/j.neuroimage.2012.01.021

Ghatan, S., De Vries, J., Pingault, J.-B., Jaddoe, V. W., Cecil, C., Felix, J. F., Rivadeneira, F., & Medina-Gomez, C. (2025). Genetic nurture: Estimating the direct genetic effects of pediatric anthropometric traits. Human Molecular Genetics, 34(20), 1744–1752. 10.1093/hmg/ddaf117

Grotzinger, A. D., Rhemtulla, M., de Vlaming, R., Ritchie, S. J., Mallard, T. T., Hill, W. D., Ip, H. F., Marioni, R. E., McIntosh, A. M., Deary, I. J., Koellinger, P. D., Harden, K. P., Nivard, M. G., & Tucker-Drob, E. M. (2019). Genomic structural equation modelling provides insights into the multivariate genetic architecture of complex traits. Nature Human Behaviour, 3(5), Article 5. 10.1038/s41562-019-0566-x

Han, L. K. M., Dinga, R., Hahn, T., Ching, C. R. K., Eyler, L. T., Aftanas, L., Aghajani, M., Aleman, A., Baune, B. T., Berger, K., Brak, I., Filho, G. B., Carballedo, A., Connolly, C. G., Couvy-Duchesne, B., Cullen, K. R., Dannlowski, U., Davey, C. G., Dima, D., … Schmaal, L. (2021). Brain aging in major depressive disorder: Results from the ENIGMA major depressive disorder working group. Molecular Psychiatry, 26(9), 5124–5139. 10.1038/s41380-020-0754-0

Hemani, G., Tilling, K., & Smith, G. D. (2017). Orienting the causal relationship between imprecisely measured traits using GWAS summary data. PLoS Genetics, 13(11), Article 11. 10.1371/journal.pgen.1007081

Hemani, G., Zheng, J., Elsworth, B., Wade, K. H., Haberland, V., Baird, D., Laurin, C., Burgess, S., Bowden, J., Langdon, R., Tan, V. Y., Yarmolinsky, J., Shihab, H. A., Timpson, N. J., Evans, D. M., Relton, C., Martin, R. M., Davey Smith, G., Gaunt, T. R., & Haycock, P. C. (2018). The MR-Base platform supports systematic causal inference across the human phenome. eLife, 7, e34408. 10.7554/eLife.34408

Hölzel, M., Rohrmoser, M., Schlee, M., Grimm, T., Harasim, T., Malamoussi, A., Gruber-Eber, A., Kremmer, E., Hiddemann, W., Bornkamm, G. W., & Eick, D. (2005). Mammalian WDR12 is a novel member of the Pes1–Bop1 complex and is required for ribosome biogenesis and cell proliferation. The Journal of Cell Biology, 170(3), 367–378. 10.1083/jcb.200501141

Jawinski, P., Forstbach, H., Kirsten, H., Beyer, F., Villringer, A., Witte, A. V., Scholz, M., Ripke, S., & Markett, S. (2025). Genome-wide analysis of brain age identifies 59 associated loci and unveils relationships with mental and physical health. Nature Aging, 5(10), 2086–2103. 10.1038/s43587-025-00962-7

Jorda, P., Jeuken, A., Lahrouchi, N., Bezzina, C. R., & Tadros, R. (2023). Assessment of multi-trait analysis of GWAS (MTAG) for discovery of novel genetic variants and mechanistic insight in common cardiovascular diseases. European Heart Journal, 44(Supplement_2), ehad655.3053. 10.1093/eurheartj/ehad655.3053

Jordà, P., Lai, Y., Jeuken, A., Lemieux Perreault, L.-P., Goulet, E., Lahrouchi, N., Nozza, A., Tanck, M. W., Guerra, P., Cadrin-Tourigny, J., De Denus, S., Bezzina, C. R., Lettre, G., Busseuil, D., Dubé, M.-P., Tardif, J.-C., & Tadros, R. (2025). Genetic analyses across cardiovascular traits: Leveraging genetic correlations to empower locus discovery and prediction in common cardiovascular diseases. Npj Genomic Medicine, 10(1), 65. 10.1038/s41525-025-00515-2

Kaufmann, T., van der Meer, D., Doan, N. T., Schwarz, E., Lund, M. J., Agartz, I., Alnæs, D., Barch, D. M., Baur-Streubel, R., Bertolino, A., Bettella, F., Beyer, M. K., Bøen, E., Borgwardt, S., Brandt, C. L., Buitelaar, J., Celius, E. G., Cervenka, S., Conzelmann, A., … Karolinska Schizophrenia Project (KaSP). (2019). Common brain disorders are associated with heritable patterns of apparent aging of the brain. Nature Neuroscience, 22(10), 1617–1623. 10.1038/s41593-019-0471-7

Kolbeinsson, A., Filippi, S., Panagakis, Y., Matthews, P. M., Elliott, P., Dehghan, A., & Tzoulaki, I. (2020). Accelerated MRI-predicted brain ageing and its associations with cardiometabolic and brain disorders. Scientific Reports, 10(1), Article 1. 10.1038/s41598-020-76518-z

Kooijman, M. N., Kruithof, C. J., Van Duijn, C. M., Duijts, L., Franco, O. H., Van IJzendoorn, M. H., De Jongste, J. C., Klaver, C. C. W., Van Der Lugt, A., Mackenbach, J. P., Moll, H. A., Peeters, R. P., Raat, H., Rings, E. H. H. M., Rivadeneira, F., Van Der Schroeff, M. P., Steegers, E. A. P., Tiemeier, H., Uitterlinden, A. G., … Jaddoe, V. W. V. (2016). The Generation R Study: Design and cohort update 2017. European Journal of Epidemiology, 31(12), 1243–1264. 10.1007/s10654-016-0224-9

Leonardsen, E. H., Vidal-Piñeiro, D., Roe, J. M., Frei, O., Shadrin, A. A., Iakunchykova, O., De Lange, A.-M. G., Kaufmann, T., Taschler, B., Smith, S. M., Andreassen, O. A., Wolfers, T., Westlye, L. T., & Wang, Y. (2023). Genetic architecture of brain age and its causal relations with brain and mental disorders. Molecular Psychiatry, 28(7), 3111–3120. 10.1038/s41380-023-02087-y

Liberzon, A., Subramanian, A., Pinchback, R., Thorvaldsdóttir, H., Tamayo, P., & Mesirov, J. P. (2011). Molecular signatures database (MSigDB) 3.0. Bioinformatics (Oxford, England), 27(12), Article 12. 10.1093/bioinformatics/btr260

Linli, Z., Feng, J., Zhao, W., & Guo, S. (2022). Associations between smoking and accelerated brain ageing. Progress in Neuro-Psychopharmacology and Biological Psychiatry, 113, 110471. 10.1016/j.pnpbp.2021.110471

Mounier, N., & Kutalik, Z. (2023). Bias correction for inverse variance weighting Mendelian randomization. Genetic Epidemiology, gepi.22522. 10.1002/gepi.22522

Paone, A., Marani, M., Fiascarelli, A., Rinaldo, S., Giardina, G., Contestabile, R., Paiardini, A., & Cutruzzolà, F. (2014). SHMT1 knockdown induces apoptosis in lung cancer cells by causing uracil misincorporation. Cell Death & Disease, 5(11), e1525–e1525. 10.1038/cddis.2014.482

Pingault, J.-B., Barkhuizen, W., Wang, B., Hannigan, L. J., Eilertsen, E. M., Corfield, E., Andreassen, O. A., Ask, H., Tesli, M., Askeland, R. B., Davey Smith, G., Stoltenberg, C., Davies, N. M., Reichborn-Kjennerud, T., Ystrom, E., & Havdahl, A. (2023). Genetic nurture versus genetic transmission of risk for ADHD traits in the Norwegian Mother, Father and Child Cohort Study. Molecular Psychiatry, 28(4), 1731–1738. 10.1038/s41380-022-01863-6

Raz, N., & Rodrigue, K. M. (2006). Differential aging of the brain: Patterns, cognitive correlates and modifiers. Neuroscience & Biobehavioral Reviews, 30(6), Article 6. 10.1016/j.neubiorev.2006.07.001

Schulz, M.-A., Siegel, N. T., & Ritter, K. (2025). Brain-age models with lower age prediction accuracy have higher sensitivity for disease detection. PLOS Biology, 23(10), e3003451. 10.1371/journal.pbio.3003451

Smith, S. M., Elliott, L. T., Alfaro-Almagro, F., McCarthy, P., Nichols, T. E., Douaud, G., & Miller, K. L. (2020). Brain aging comprises many modes of structural and functional change with distinct genetic and biophysical associations. eLife, 9, e52677. 10.7554/eLife.52677

Soumya Kumari, L. K., & Sundarrajan, R. (2024). A review on brain age prediction models. Brain Research, 1823, 148668. 10.1016/j.brainres.2023.148668

Tubbs, J. D., Porsch, R. M., Cherny, S. S., & Sham, P. C. (2020). The Genes We Inherit and Those We Don’t: Maternal Genetic Nurture and Child BMI Trajectories. Behavior Genetics, 50(5), 310–319. 10.1007/s10519-020-10008-w

Vidal-Pineiro, D., Wang, Y., Krogsrud, S. K., Amlien, I. K., Baaré, W. F., Bartres-Faz, D., Bertram, L., Brandmaier, A. M., Drevon, C. A., Düzel, S., Ebmeier, K., Henson, R. N., Junqué, C., Kievit, R. A., Kühn, S., Leonardsen, E., Lindenberger, U., Madsen, K. S., Magnussen, F., … Fjell, A. (2021). Individual variations in ‘brain age’ relate to early-life factors more than to longitudinal brain change. eLife, 10, e69995. 10.7554/eLife.69995

Wen, J., Tian, Y. E., Skampardoni, I., Yang, Z., Cui, Y., Anagnostakis, F., Mamourian, E., Zhao, B., Toga, A. W., Zalesky, A., & Davatzikos, C. (2024). The genetic architecture of biological age in nine human organ systems. Nature Aging, 4(9), 1290–1307. 10.1038/s43587-024-00662-8

Xu, H., Toikumo, S., Crist, R. C., Glogowska, K., Jinwala, Z., Deak, J. D., Justice, A. C., Gelernter, J., Johnson, E. C., Kranzler, H. R., & Kember, R. L. (2023). Identifying genetic loci and phenomic associations of substance use traits: A multi-trait analysis of GWAS (MTAG) study. Addiction, 118(10), 1942–1952. 10.1111/add.16229

Zettergren, A., Seidu, N., Najar, J., Marseglia, A., Dartora, C., Blennow, K., Zetterberg, H., Waern, M., Kern, S., Westman, E., & Skoog, I. (2024). Polygenic scores for longevity and cognitive performance are associated with predicted brain age in 70-year-olds from the general population. Cerebral Circulation - Cognition and Behavior, 6, 100306. 10.1016/j.cccb.2024.100306

Zhang, X., Wang, J., Zhang, Z., & Ye, K. (2024). Tau in neurodegenerative diseases: Molecular mechanisms, biomarkers, and therapeutic strategies. Translational Neurodegeneration, 13(1), 40. 10.1186/s40035-024-00429-6

Zheng, Z., Liu, S., Sidorenko, J., Wang, Y., Lin, T., Yengo, L., Turley, P., Ani, A., Wang, R., Nolte, I. M., Snieder, H., LifeLines Cohort Study, Aguirre-Gamboa, R., Deelen, P., Franke, L., Kuivenhoven, J. A., Lopera Maya, E. A., Sanna, S., Swertz, M. A., … Zeng, J. (2024). Leveraging functional genomic annotations and genome coverage to improve polygenic prediction of complex traits within and between ancestries. Nature Genetics, 56(5), 767–777. 10.1038/s41588-024-01704-y

