## Supplementary Material for "Shared genetic architecture of brain age gap across 30 cohorts worldwide"

### Supplementary Materials

**Supplementary Methods** 3
SM Methods 1.1 $BAG_{Han}$ GWAS 3
Rationale 3
$BAG_{Han}$ GWAS: Samples 4
$BAG_{Han}$ GWAS: Brain imaging and quality control 5
$BAG_{Han}$ GWAS: Phenotype construction 5
$BAG_{Han}$ GWAS: Genotype imputation and quality control 6
$BAG_{Han}$ GWAS: Genome-wide association analyses 6
$BAG_{Han}$ GWAS: Meta-analysis of GWAS results 7
$BAG_{Han}$ GWAS: SNP heritability and genetic correlations 7
$BAG_{Han}$ GWAS: Genomic annotation 8
$BAG_{Han}$ GWAS: Polygenic scores in UKBB 8
SM Methods 1.2 BAG factor post-GWAS analyses 8
Genomic loci and functional annotation 8
Gene functional annotation 9
MAGMA gene-based, gene-set and tissue expression analyses 9
Polygenic score covariates **9**
**Supplementary Results** 11
SM Results 2.1 $BAG_{Han}$ GWAS 11
$BAG_{Han}$ GWAS: GWAS meta-analysis 11
$BAG_{Han}$ GWAS: Genomic annotation 12
$BAG_{Han}$ GWAS: Genetic correlations 13
$BAG_{Han}$ GWAS: Polygenic scores in UKBB hold-out sample 14
SM Results 2.2 Two-factor solution 15
SM Results 2.3 MR sensitivity analyses 15
**Supplementary Figures** 17
**Figure S1**. Manhattan plot of $BAG_{Han}$ genome-wide association meta-analysis 17
**Figure S2.** Genetic correlations of $BAG_{Han}$ with health outcomes 18
**Figure S3**. Two-factor model of brain age gap 19

**Figure S4**. Gene expression heatmap v8 54 tissue types using average expression per label (log2 transformed) 20

**Figure S5**. Differentially expressed genes using GTEx v8 54 tissue types 22

**Figure S6**. GWAS catalog reported genes for the brain age gap factor 23

**Figure S7**. Mendelian randomisation results: Effect of 33 health outcomes on seven distinct brain age gaps 24

**Figure S8.** Mendelian randomisation results: Effect of seven distinct brain age gaps on 33 health outcomes 26

**Figure S9**. Observed and bias-corrected Mendelian randomisation effects for the association between health outcomes and brain age gap 28

**Figure S10**. PheWAS results for seven polygenic scores derived from BAG factor and six individual GWASs in the UK Biobank cohort 29

### Supplementary Methods

#### SM Methods 1.1 $BAG_{Han}$ GWAS

##### Rationale

In recent years, a growing number of brain age prediction models have been developed, based on various neuroimaging modalities, machine learning algorithms, training samples and age ranges (Kumari & Sundarrajan, 2024). The performance of these models has been typically evaluated using two key metrics: the strength of the relationship between predicted age and chronological age (using the correlation coefficient, *r*) and the average absolute difference between predicted age and chronological age (quantified by the mean absolute error [MAE]). While a well-performing model is usually characterised by a large correlation coefficient and a small MAE, it has been recently noted that a high accuracy in estimating chronological age does not necessarily equate to clinical utility (Dörfel et al., 2025; Schulz et al., 2025). For example, Bashyam et al. (2020) found that a moderately fit model (MAE = 5.92), in comparison to a tight (MAE = 3.70) or a loose model (MAE = 7.65), provided brain age estimates that were better able to identify individuals with mild cognitive impairment, Alzheimer's disease, major depression, and schizophrenia.

Given the lack of a clear consensus on whether a tight, moderate, or loose model fit is best for predicting disease-relevant information, we aimed to include brain age GWASs that pertained to each model performance category. As such, we selected $BAG_{Leonardsen,}$ $BAG_{Smith.}$, $BAG_{Jawinski}$, $BAG_{Wen}$, $BAG_{Kaufmann}$, and $BAG_{Han}$ GWASs, which together covered tight, moderate, and loose-fitting models (MAE from 2.9 to 6.8 years). As a GWAS for $BAG_{Han}$ was not yet available (our 'loose' model), we conducted one as part of the present study. In addition to representing the looser model fit category, our GWAS integrated 29 cohorts worldwide, thereby increasing sample size and global diversity compared with prior BAG GWASs, which were either solely based on the UKBB or, at most, included one other cohort.

#### BAG$_{Han}$ GWAS: Samples

Twenty-nine cohorts took part in the GWAS, including 27 cohorts from the Enhancing Neuroimaging Genetics through Meta-analysis (ENIGMA) consortium, UK Biobank, and the Avon Longitudinal Study of Parents and Children (ALSPAC). Cohort characteristics and descriptives can be found in **Tables S1-S2**. In total, data from 60,810 individuals were included ($N = 28{,}386$ males; $N = 32{,}424$ females), of which 60,735 had genetic data and were included in the final sample of this GWAS. All participating cohorts obtained ethical approval from their respective institutional review boards or ethics committees. Participants were eligible for inclusion in the study if they met the following criteria:

1) Age 18 or older
2) Genetic and structural MRI data were available
3) Key covariates were available (i.e., age, sex, intracranial volume, and genetic principal components)
4) Passed basic quality control (QC) filters (e.g., no discrepancy in genetic and assigned sex, no individuals with heterozygosity rates ±3 SD from the sample mean)
5) No BAG outlier (±5 IQR from the sample mean)

Note that to preserve a larger sample size, related individuals were *not* excluded. Instead, we used RareMetalWorker (Feng et al., 2014), which controls for related individuals and hidden population structure via a genomic relationship matrix estimated from genotype data. Furthermore, we did not exclude individuals based on disease status. Instead, disease status was included as a covariate in the GWAS model. Although our analysis included individuals from diverse ancestral backgrounds, the majority were of European ancestry (**Table S1**). Only cohorts with a minimum of 50 individuals were included to ensure we obtain stable predictions in genetic analyses.

**$BAG_{Han}$ GWAS: Brain imaging and quality control**

Structural T1-weighted scans were processed using FreeSurfer v. 5.1 to 7.4 (Fischl, 2012) resulting in 76 region-of-interest (ROI) measures based on the Desikan/Killiany atlas (Desikan et al., 2006): 34 cortical thickness measures, 34 cortical surface area measures, 7 subcortical volumes, and 1 lateral ventricle volume, calculated as the mean across the left and right hemispheres, along with total intracranial volume (i.e., 77 features in total). All cohorts (apart from the UKBB) followed the ENIGMA quality control protocol detailed here: https://enigma.ini.usc.edu/protocols/imaging-protocols/, which involved visually inspecting cortical and subcortical segmentations and identifying and (if deemed necessary) removing outliers. Further details on image acquisition parameters and the number of subjects removed due to failing quality control may be found in **Table S3**.

**$BAG_{Han}$ GWAS: Phenotype construction**

The 77 features were used as input to ENIGMA's ridge regression model, developed by Han et al. (2021). The $BAG_{Han}$ model was trained on structural brain measures from 952 healthy males and 1,236 healthy females, aged 18-75 years, drawn from the ENIGMA Major Depressive Disorder group (see **Table 1**). The parameters from the trained model were applied to our samples (separately for males and females), to generate brain-predicted age estimates for each individual. BAG was calculated by subtracting the chronological age of each individual from their brain-predicted age. To evaluate the model's performance in our samples, we calculated the mean absolute error (MAE) and the Pearson correlation coefficient (*r*) between predicted brain age and chronological age. For further details related to model development, see Han et al. (2021).

#### BAG$_{Han}$ GWAS: Genotype imputation and quality control

All cohorts imputed their genotypes using the 1000 Genomes Project (The 1000 Genomes Project Consortium, 2010), apart from i-Share that used the Haplotype Reference Consortium (the Haplotype Reference Consortium, 2016), and UKBB that used the Haplotype Reference Consortium, UK10K, and 1000 Genomes Project Phase 3). Quality control steps included filtering for minor allele frequency, individual missingness, genotype call rate, and Hardy-Weinberg equilibrium. For concrete quality control thresholds and cohort-specific exclusion criteria (e.g., gender mismatches, minimal or excessive heterozygosity), see **Table S4**.

#### BAG$_{Han}$ GWAS: Genome-wide association analyses

We used genetic association analyses to estimate additive effects of genetic variants on BAG, including age, age$^2$, sex, total intracranial volume, and 4 genetic principal components (PCs) as covariates. When applicable, analyses were also adjusted for case-control status and imaging site. We applied a linear mixed model using the RareMetalWorker software (RMW; Feng et al., 2014). RMW handles related individuals and hidden population structure via a genomic relationship matrix estimated from genotype data. For the UK Biobank cohort, we used Bayesian linear mixed model (BOLT-LMM; Loh et al., 2015) software as it is more computationally efficient with large sample sizes. For the UK Biobank GWAS, in addition to the core covariates, we included genotyping array to control for array effects, and increased the number of genetic PCs from four to 10 to better account for population structure in this larger sample. Lastly, adapting to cohort-unique requirements, we used REGENIE (Mbatchou et al.,

2021) for i-Share and PLINK 2.0 (Chang et al., 2015) for ALSPAC. The analyses were kept as similar as possible across cohorts to minimise potential sources of heterogeneity. The Manhattan and quantile–quantile (QQ) plots were visually inspected for each cohort to ensure genetic analyses were performed appropriately.

##### $BAG_{Han}$ GWAS: Meta-analysis of GWAS results

The resulting summary statistics from each cohort were meta-analysed via METAL using fixed effects meta-analysis (Kavvoura & Ioannidis, 2008), which assumes that the true effect of each risk allele is consistent across datasets and that any study-specific findings are due to sampling variation (Evangelou & Ioannidis, 2013). The meta-analysis was conducted in two stages. We first meta-analysed 28 smaller cohorts, each with $n < 2{,}000$ (total $n = 14{,}413$ across all 28 cohorts). Subsequently, these results were combined with the UK Biobank data ($n = 46{,}322$) in a second-stage meta-analysis. The two-stage approach was used to prevent extreme sample size imbalances from disproportionately driving the meta-analysis results, as previously done in Grasby et al. (2020). In both stages of the meta-analysis, sample size weighting was applied. Heterogeneity across cohorts (the possibility that the true effect sizes between studies are different) was assessed using the $I^2$ statistic. Only SNPs with MAF > 0.1 and INFO score > 0.6 were included in the meta-analysis. Variants reaching genome-wide significance ($p < 5e\text{-}8$) were compared against previously reported variants from five prior BAG GWASs (Jawinski et al., 2025; Kaufmann et al., 2019; Leonardsen et al., 2023; Smith et al., 2020; Wen et al., 2024).

##### $BAG_{Han}$ GWAS: SNP heritability and genetic correlations

We used linkage disequilibrium (LD) score regression (Bulik-Sullivan et al., 2015; Bulik-Sullivan et al., 2015) to obtain SNP-based heritability of $BAG_{Han}$ and to perform genetic correlations analyses with 33 psychiatric, cardiometabolic, behavioural, and aging-related traits.

**$BAG_{Han}$ GWAS: Genomic annotation**

We used FUMA (Functional Mapping and Annotation v.1.5.2; Watanabe et al., 2017) to annotate, prioritise, and visualize GWAS results. Specifically, we used *SNP2GENE* function to obtain functional annotation for all SNPs and *GENE2FUNC* function to annotate genes in a biological context. SNPs were mapped to genes using (1) positional mapping (±10 kb), (2) eQTL mapping with GTEx v8, PsychENCODE, and BRAINEAC data repositories, (3) chromatin interaction mapping using enhancer–promoter and Hi-C datasets, and (4) MAGMA gene-based analysis.

**$BAG_{Han}$ GWAS: Polygenic scores in UKBB**

As a validation step, we constructed a polygenic score using SBayesRC (Zheng et al., 2024) based on $BAG_{Han}$ summary statistics and tested their association with phenotypic BAG in a hold-out UKBB sample stratified by ancestry group (White, Asian, Black, Mixed; UKBB Data-Field 21000; $n$=3266; **Figure 1**). SBayesRC integrates functional genomic annotations alongside high-density SNP data (>7M variants) to improve polygenic prediction. This analysis allowed us to assess the proportion of variance in BAG explained by the polygenic score.

#### SM Methods 1.2 BAG factor post-GWAS analyses

**Genomic loci and functional annotation.** To annotate, prioritise and visualise GWAS results, we used FUMA v1.5.2 (Watanabe et al., 2017) *SNP2GENE* pipeline. Independent genome-wide significant variants were defined as those with $p < 5e\text{-}8$ and in low linkage disequilibrium (LD) with each other ($r^2 < 0.1$). LD blocks around independent significant SNPs were merged into a single genomic risk locus using a 500 kb distance. Genetic variants were mapped to genes using three mapping techniques: positional (±10 kb window size), expression quantitative trait loci (eQTL), and chromatin interaction mapping. Mapping was based on PsychENCODE, BRAINEAC, GTEx v8, FANTOM5, and Hi-C datasets (Akbarian et al., 2015; Kawaji et al., 2017; Ramasamy et al., 2014; Schmitt et al., 2016; The GTEx Consortium et al., 2020).

**Gene functional annotation**. Candidate genes identified via at least one of the three mapping techniques were used as input to FUMA's *GENE2FUNC* pipeline, which provided a biological context for the identified genes. Tissue-specific gene expression was visualised using heatmaps based on GTEx v8 data for 54 specific and 30 general tissue types. Differentially expressed genes were identified by testing for enrichment of prioritised genes against background gene sets. Gene-set enrichment analysis was performed to test if selected genes are overrepresented in any of the pre-defined gene sets, including MSigDB (Liberzon et al., 2011), WikiPathways (Slenter et al., 2018), and GWAS catalog-reported genes (Buniello et al., 2019).

**MAGMA gene-based, gene-set and tissue expression analyses.** Gene-based association analysis was performed using MAGMA v1.08. SNPs were assigned to 18,781 protein-coding genes using a ±10 kb window. SNP-level *p*-values were combined to obtain gene-level *p*-values. Gene-set analysis was performed for curated gene sets and GO terms obtained from the MsigDB database (Liberzon et al., 2011).

**Polygenic score covariates.** In UKBB, sex, age, genotyping array, and 10 genetic principal components were controlled for, with age$^2$, total intracranial volume, and scanner site only included in the validation analyses. In Generation R, polygenic scores were standardised and residualised for the first five genetic principal components; age and sex were included as covariates.

### Supplementary Results

#### SM Results 2.1 $BAG_{Han}$ GWAS

##### $BAG_{Han}$ GWAS: GWAS meta-analysis

We performed a genome-wide association meta-analysis of $BAG_{Han}$ across 29 cohorts (total $N = 60{,}735$; mean age=37.6 years [range=18-75]; 53% female). We identified 32 lead SNPs across 29 genomic risk loci reaching genome-wide significance ($p < 5e^{-8}$) using LD clumping at $r^2 < 0.1$ with a 500 kb window, with generally low to moderate heterogeneity across cohorts, which was largely not statistically significant (HetP > 0.05 for most SNPs; **Table S5; Figure S1**). The GWAS yielded a genomic control lambda (λGC) of 1.22 and an LD score regression intercept of 1.02 (SE=0.01), indicating inflation driven primarily by polygenicity. The identified SNPs mapped to 718 genes (**Table S6**). Fifty-seven of these genes (e.g., *DPYSL5*, *PSENEN*, *MLH1*) were identified using all four mapping techniques (positional, eQTL, chromatin interaction, and MAGMA), providing the strongest support for the involvement of these genes in brain age (**Table S7**). The SNP-based heritability ($h^2_{SNP}$) of the brain age gap was estimated at 0.21 (SE = 0.01). In the total sample, performance metrics showed an average MAE of 9.08 years (SD=5.98) and moderate correlations between brain-predicted age and chronological age ($r_{female} = 0.49$, $r_{male} = 0.52$). For individual performance metrics of all 29 cohorts included in $BAG_{Han}$ meta-analysis see **Table S8**.

Out of 32 identified variants, only one had been identified in a prior BAG GWAS (rs12146713; Jawinski et al., 2025), while seven additional variants reached suggestive significance ($p$<5e-5; Jawinski et al., 2025; Kaufmann et al., 2019; Wen et al., 2024), indicating potentially novel SNPs. We note that this refers to direct (SNP-specific) replication, and not all SNPs were present in all GWASs (**Table S5**).

**Figure S1.** Manhattan plot of $BAG_{Han}$ genome-wide association meta-analysis

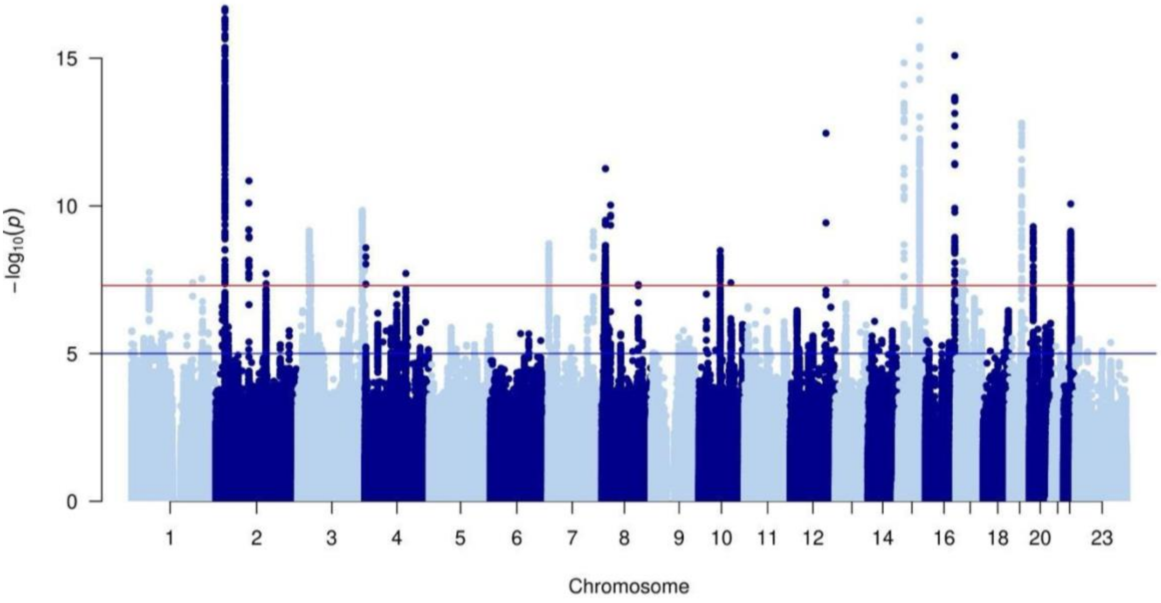


*Note.* The y-axis depicts $-\log_{10}(p)$ values for genetic variants associated with BAG. The genome-wide significance threshold is denoted by the horizontal red line at $p$ = 5e-8. The horizontal blue line denotes suggestive significance at $p$ = 5e-5.

#### $BAG_{Han}$ GWAS: Genomic annotation

A cluster of genes involved in neuronal structure, synaptic function and metabolism (e.g., *C4orf48*, *NAT8L*, *NEFM*, *APLP1*, *SNAP25*; (Endele et al., 2011; Kharel et al., 2023; Muñoz-Lasso et al., 2020; Schilling et al., 2017; Tafoya et al., 2006) showed higher expression in the brain compared to peripheral tissues (**Table S9)**. However, in differential expression analysis, no differentially expressed gene sets remained significantly enriched after Bonferroni correction (**Table S10**), suggesting no tissue-specific expression patterns. GWAS Catalog analysis revealed that mapped genes had been previously associated with brain-related morphology (i.e., brain region volumes, cortical thickness/surface area, white matter microstructure, cerebellar volume), cognitive and psychiatric traits (e.g., educational attainment, cognitive performance, schizophrenia, bipolar disorder, neuroticism), and other health-related measures (e.g., body mass index, waist-hip ratio, lipids, blood counts, kidney function; **Table S11**).

##### $BAG_{Han}$ GWAS: Genetic correlations

$BAG_{Han}$ genetically correlated with five out of 33 tested traits (cigarettes per day, diastolic blood pressure, resting heart rate, Parkinson's disease, and longevity), of which none survived FDR correction (**Figure S2**; **Table S12**).

**Figure S2.** Genetic correlations of $BAG_{Han}$ with 33 health outcomes

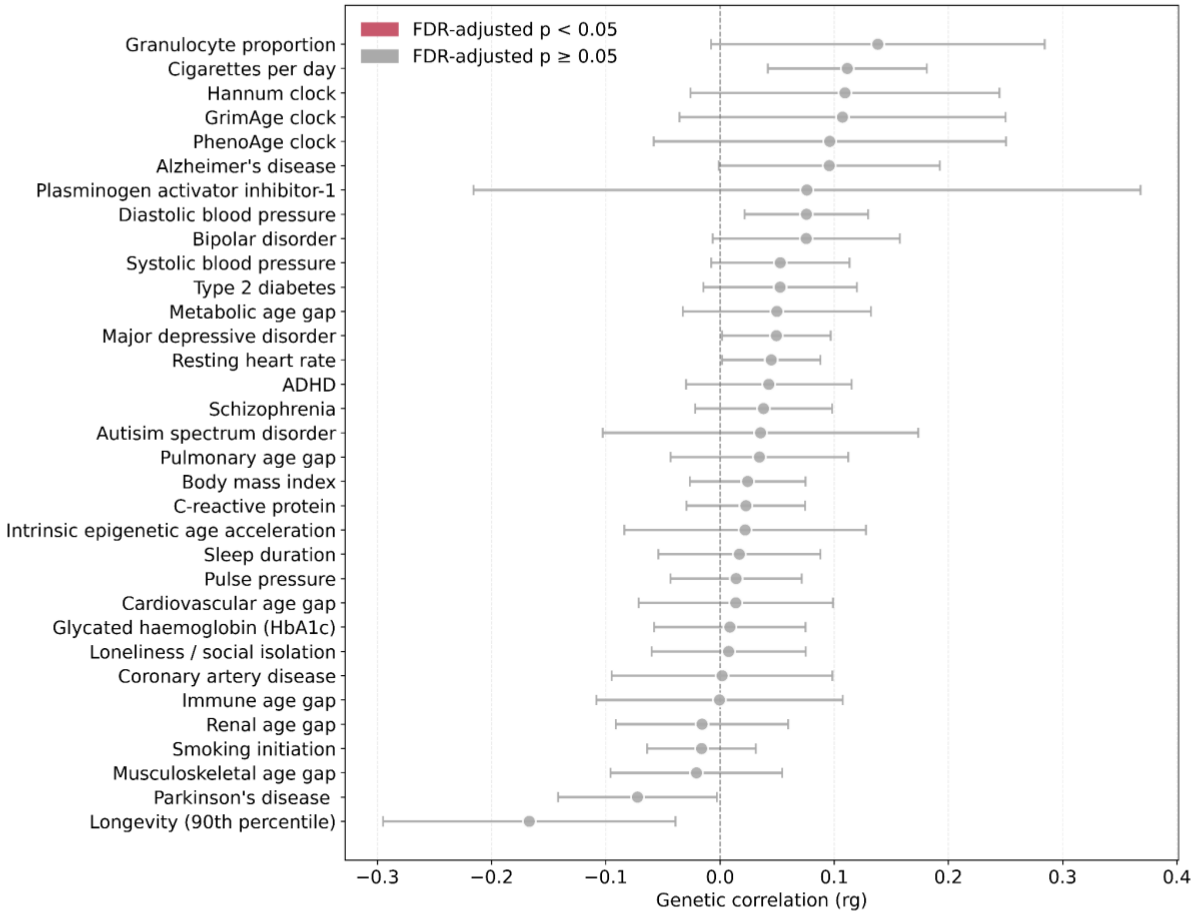


*Note.* ADHD = attention deficit hyperactivity disorder.

**BAG$_{Han}$ GWAS: Polygenic scores in UKBB hold-out sample**

Polygenic scores based on BAG$_{Han}$ explained the largest proportion of variance in brain age gap in European ancestry individuals ($R^2$=8.3%; *n*=1,738; see Table below). In other ancestry groups, the variance explained was substantially smaller ($R^2$=1.2-2.7%), which suggests that the polygenic score may not generalise well to non-European ancestries.

**Table**. Associations of BAG$_{Han}$ polygenic score with phenotypic BAG in a hold-out UK Biobank sample

| **Ancestry** | $R^2$ | **rho** | **t-value** | **n** | ***p*** |
|---|---|---|---|---|---|
| European (EUR) | 0.083 | 0.288 | 12.533 | 1738 | 3.2E-34 |
| African (AFR) | 0.012 | 0.111 | 1.986 | 316 | 0.054 |
| Central/South Asian (CSA) | 0.027 | 0.164 | 4.018 | 586 | 8.5E-05 |
| East-Asian (EAS) | 0.016 | 0.125 | 2.071 | 271 | 0.046 |

*Note.* This table presents a subset of the results available in Table S28. $R^2$ = proportion of variance in the BAG phenotype explained by the polygenic score, computed as the square of the partial correlation coefficient ($rho^2$). rho = partial product-moment correlation coefficient between the polygenic score and the BAG phenotype, adjusted for sex, age, $age^2$, total intracranial volume, scanner site, genotyping array, and 10 genetic principal components. t-value = t-statistic used to assess the significance of the partial correlation between the polygenic score and the BAG phenotype. n = sample size of the target dataset used in the association analysis.

#### SM Results 2.2 Two-factor solution

A 2-factor model improved model fit ($\chi2[8]=32.64$, $p=7.14^{-05}$, CFI=0.98, SRMR=0.06, AIC=58.64; **Figure S3**), explaining 77% of the variance. $BAG_{Leondardsen/Jawinski/Wen/Smith}$ loaded onto the first factor (all standardised loadings > 0.78), while $BAG_{Han/Kaufmann}$ loaded onto the second factor (loadings > 0.73). Despite this, the 1-factor model was retained based on good model fit, high correlation between factors ($r = 0.72$), and the study's focus on shared genetic variance.

#### SM Results 2.3 MR sensitivity analyses

Estimates from MR sensitivity analyses (weighted median and MR-Egger) were directionally concordant with primary inverse variance weighted (IVW) estimates for diastolic blood pressure, longevity, and smoking initiation, but not for systolic blood pressure (**Table S21**). MR-Egger intercept tests showed no evidence of horizontal pleiotropy (all intercepts were close to zero and non-significant [$p > 0.05$]; see **Table S21**). However, Cochran's Q statistic revealed significant heterogeneity among the genetic variants for systolic blood pressure, diastolic blood pressure, and longevity, suggesting potential violation of the instrumental variable assumptions. No significant heterogeneity was detected for smoking initiation (**Table S21**). After removing potentially invalid SNPs via Steiger filtering, we observed consistent results (i.e., consistent in direction of effect and significance level with the primary estimate; **Table S23**).

In reverse MR analyses (BAG on trait), MR sensitivity estimates were directionally consistent with the primary IVW estimate for autism spectrum disorder and sleep duration, but not for smoking initiation (**Table S22**). While MR-Egger intercept tests showed no evidence of horizontal pleiotropy, the Cochran's Q statistic indicated evidence of heterogeneity for smoking initiation and sleep duration (**Table S22)**. When performing Steiger filtering, all instruments were retained for the three traits, indicating alignment with the assumed causal direction (**Table S23**).

Forward MRlap analyses suggested that effect estimates for blood pressure traits and longevity were unlikely to be meaningfully biased upwards due to sample overlap between the exposure and outcome data or winner's curse, or biases due to the use of weak instruments. On the other hand, the association with smoking initiation appeared to be more impacted by these biases (**Figure S10**; **Table S24**). MRlap analyses in the reverse direction indicated that effect estimates for autism spectrum disorder, smoking initiation, and sleep duration were also unlikely to be substantially biased upwards due to sample overlap (**Figure S10**; **Table S24**).

### Supplementary Figures

**Figure S1**. Manhattan plot of $BAG_{Han}$ genome-wide association meta-analysis

[Figure S1 can be found in Supplementary Results].

**Figure S2.** Genetic correlations of $BAG_{Han}$ with health outcomes

[Figure S2 can be found in Supplementary Results].

**Figure S3**. Two-factor model of brain age gap

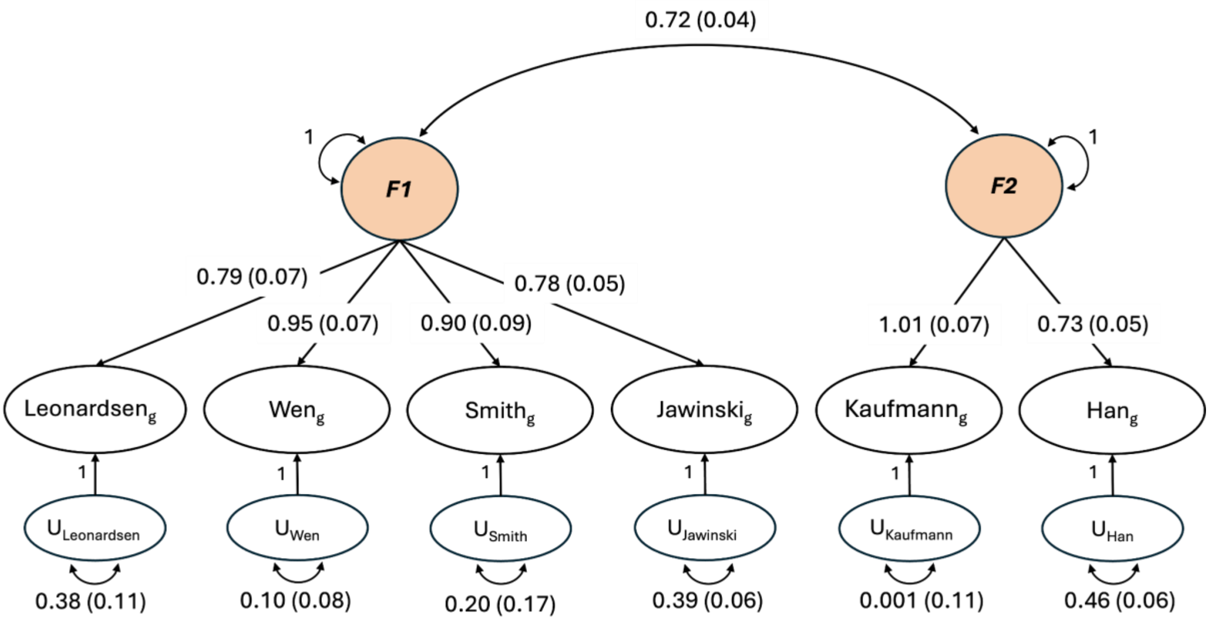


*Note*. Model fit statistics for the 2-factor confirmatory factor analysis: $\chi^2$(df) = 32.64, $p < .001$; AIC = 58.64; CFI = 0.98; SRMR = 0.06. F = latent factors; U = unique variances (error terms); single-headed arrows represent standardised factor loadings; double-headed arrows represent correlations between latent factors and covariances between residuals. Variances of latent factors were fixed to 1.

**Figure S4**. Gene expression heatmap v8 54 tissue types using average expression per label (log2 transformed)

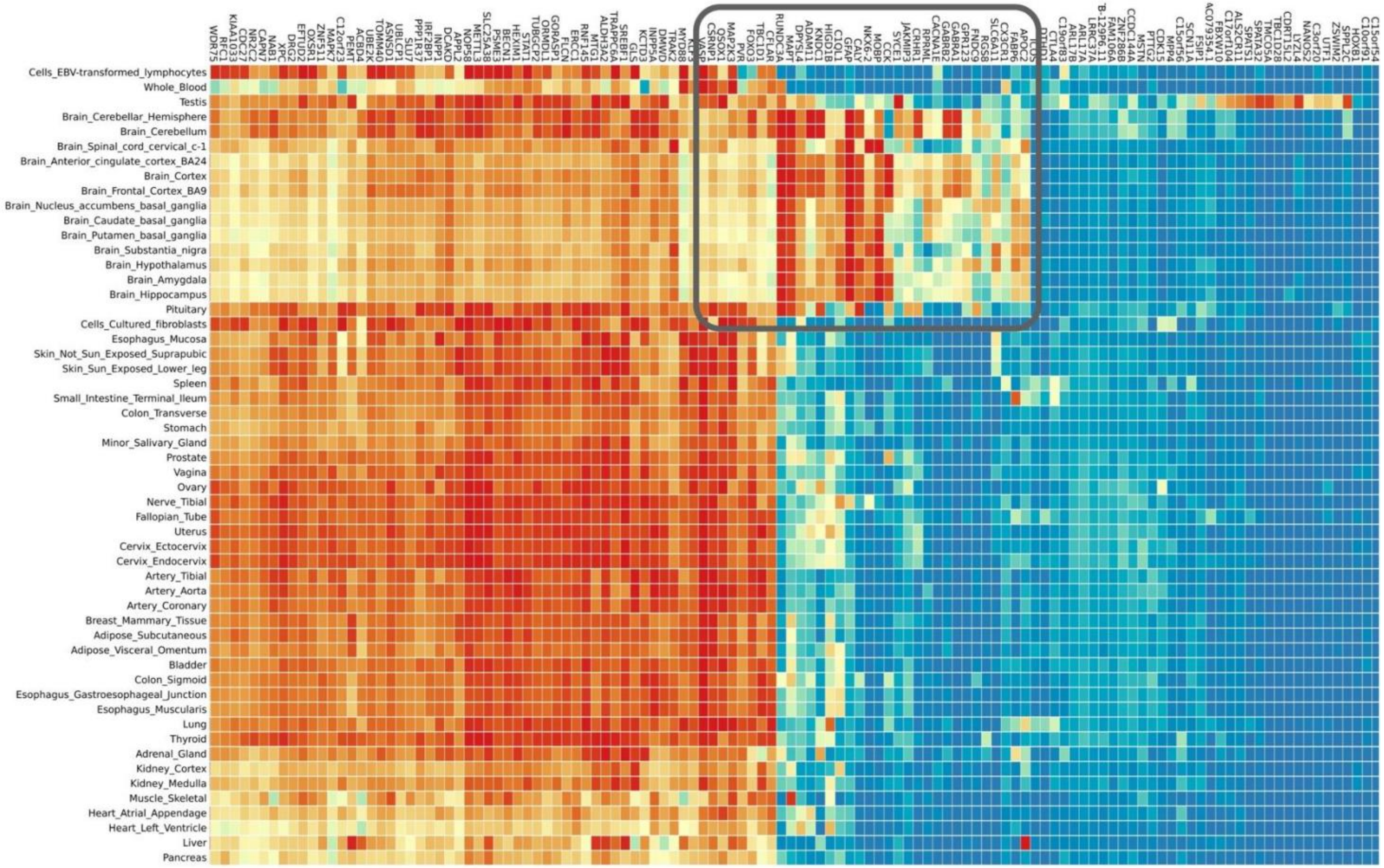

*Note*. The average of $log_2$-transformed expression values within each tissue is depicted. Darker red indicates higher expression of that gene, whereas darker blue indicates lower expression. The figure has been cropped to highlight higher expression in brain tissue compared to peripheral tissues (grey square). For the full set of results, see Table S16.

**Figure S5**. Differentially expressed genes using GTEx v8 54 tissue types

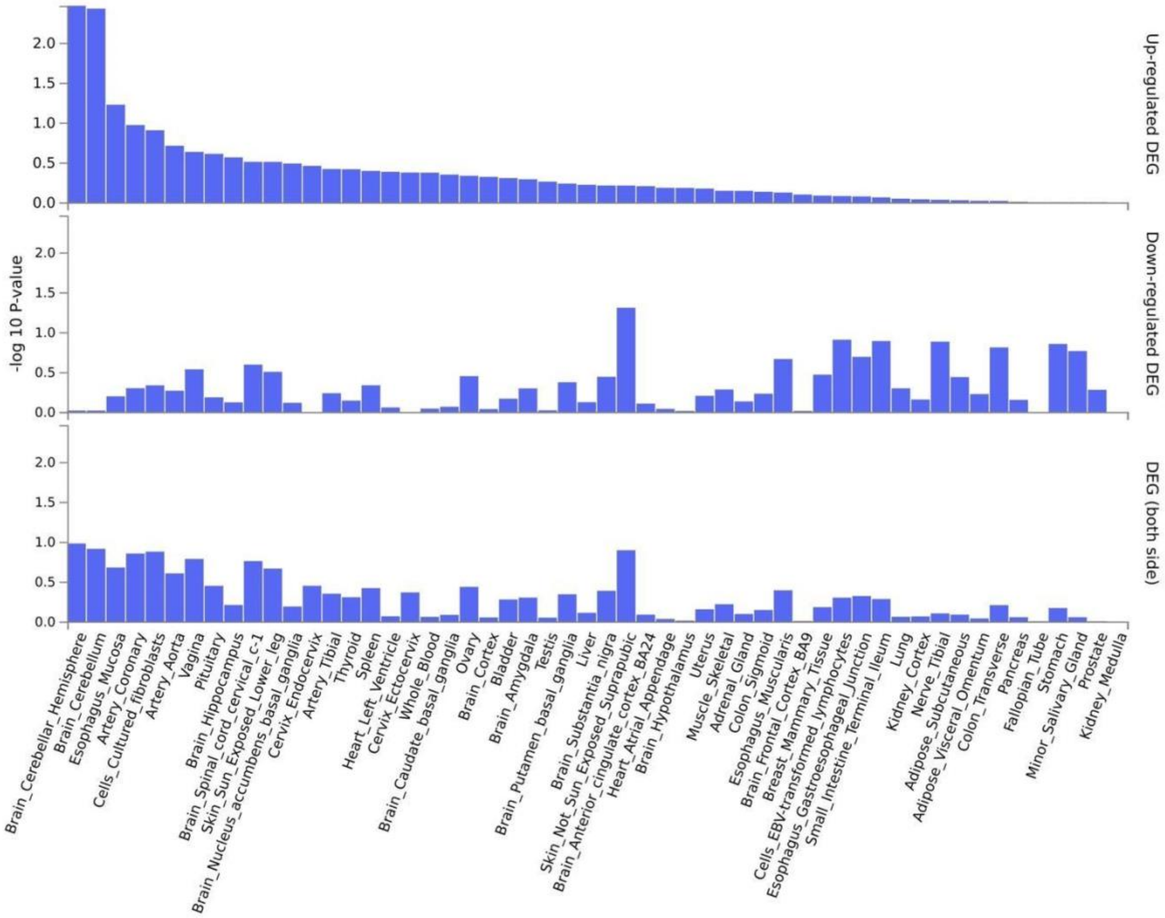

**Figure S6**. GWAS catalog reported genes for the brain age gap factor

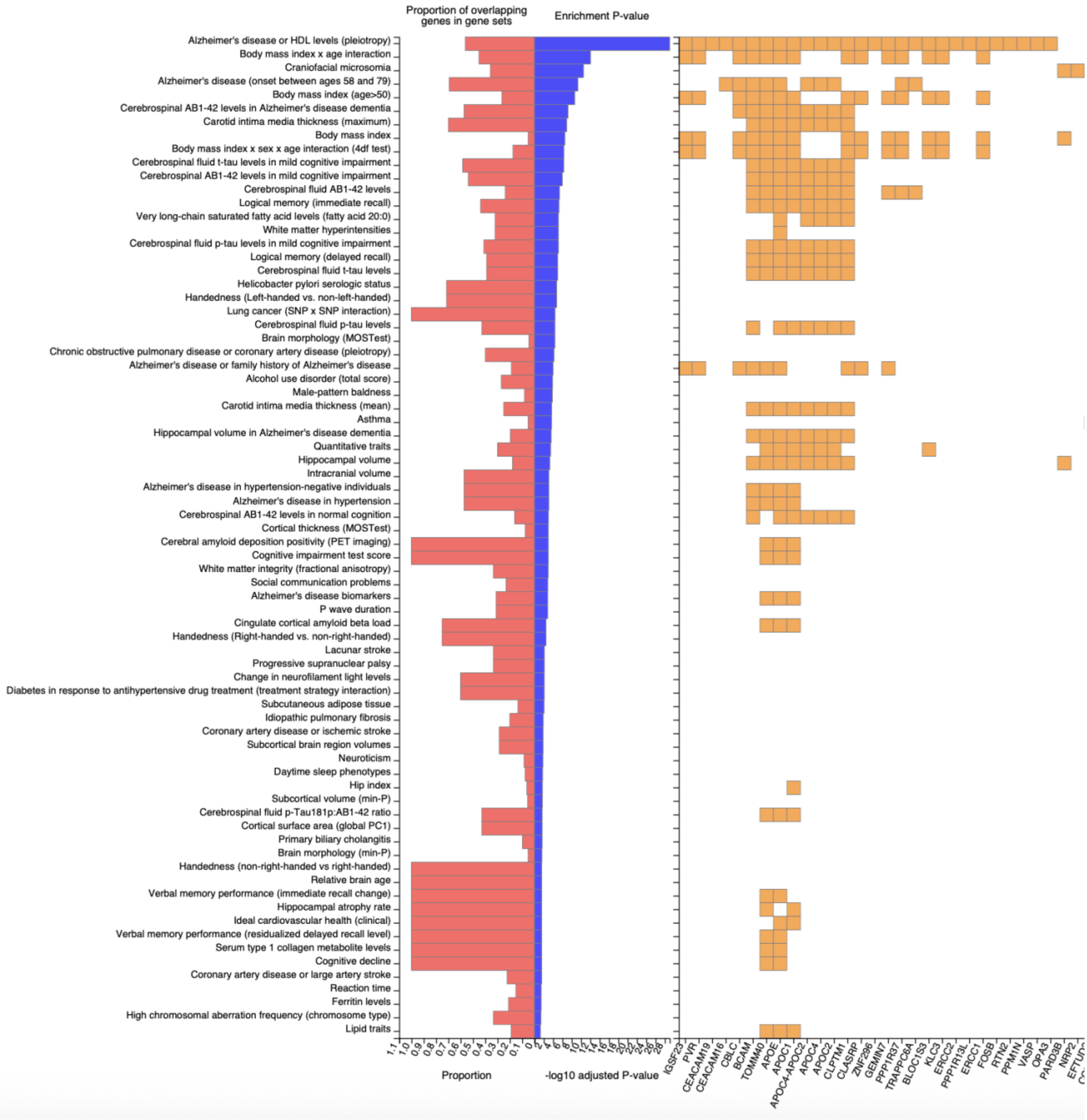


*Note*. Gene list is cropped. For the full set of genes, see Table S19.

**Figure S7**. Mendelian randomisation results: Effect of 33 health outcomes on seven distinct brain age gaps

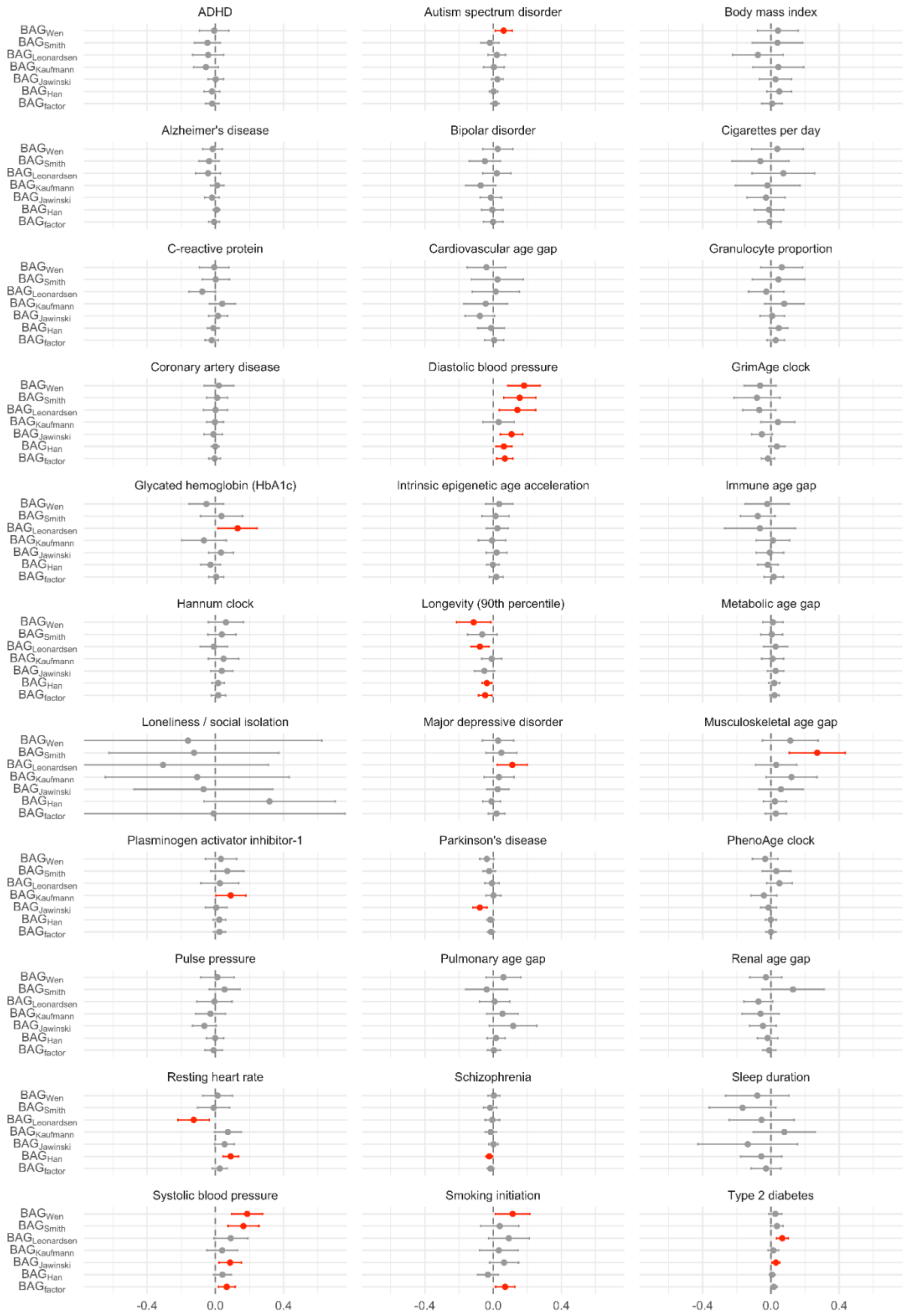

*Note*. Inverse variance weighted estimates are plotted. For sensitivity analysis estimates, see Table S21. ADHD = attention deficit hyperactivity disorder. BAG = brain age gap.

**Figure S8.** Mendelian randomisation results: Effect of seven distinct brain age gaps on 33 health outcomes

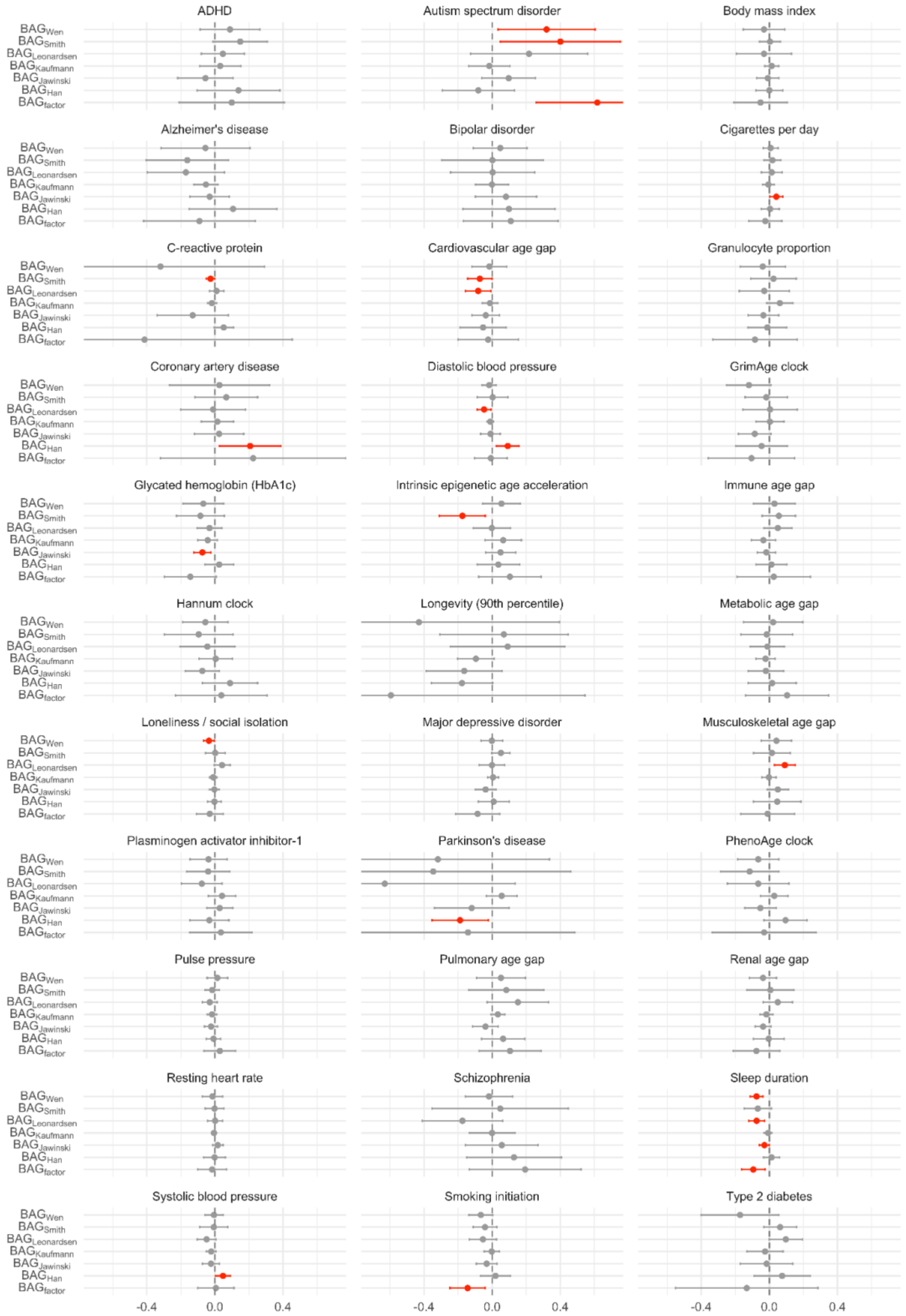

*Note*. Inverse variance weighted estimates are plotted. For sensitivity analysis estimates, see Table S22. ADHD = attention deficit hyperactivity disorder. BAG = brain age gap.

**Figure S9**. Observed and bias-corrected Mendelian randomisation effects for the association between health outcomes and brain age gap

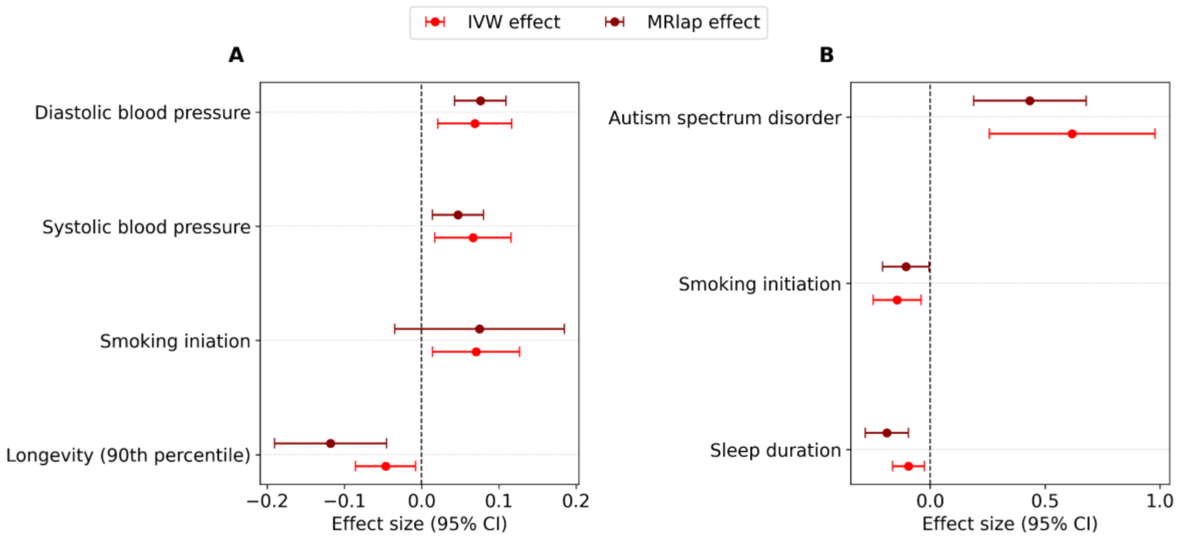


*Note*. (**A**) Displays the effect of health outcomes on brain age gap factor. (**B**) Displays the effect of brain age gap factor on health outcomes. IVW = observed inverse variance weighted estimate from primary MR analysis. MRlap = corrected estimate from MRlap sensitivity analysis. MR = Mendelian randomisation.

**Figure S10**. PheWAS results for seven polygenic scores derived from BAG factor and six individual GWASs in the UK Biobank cohort

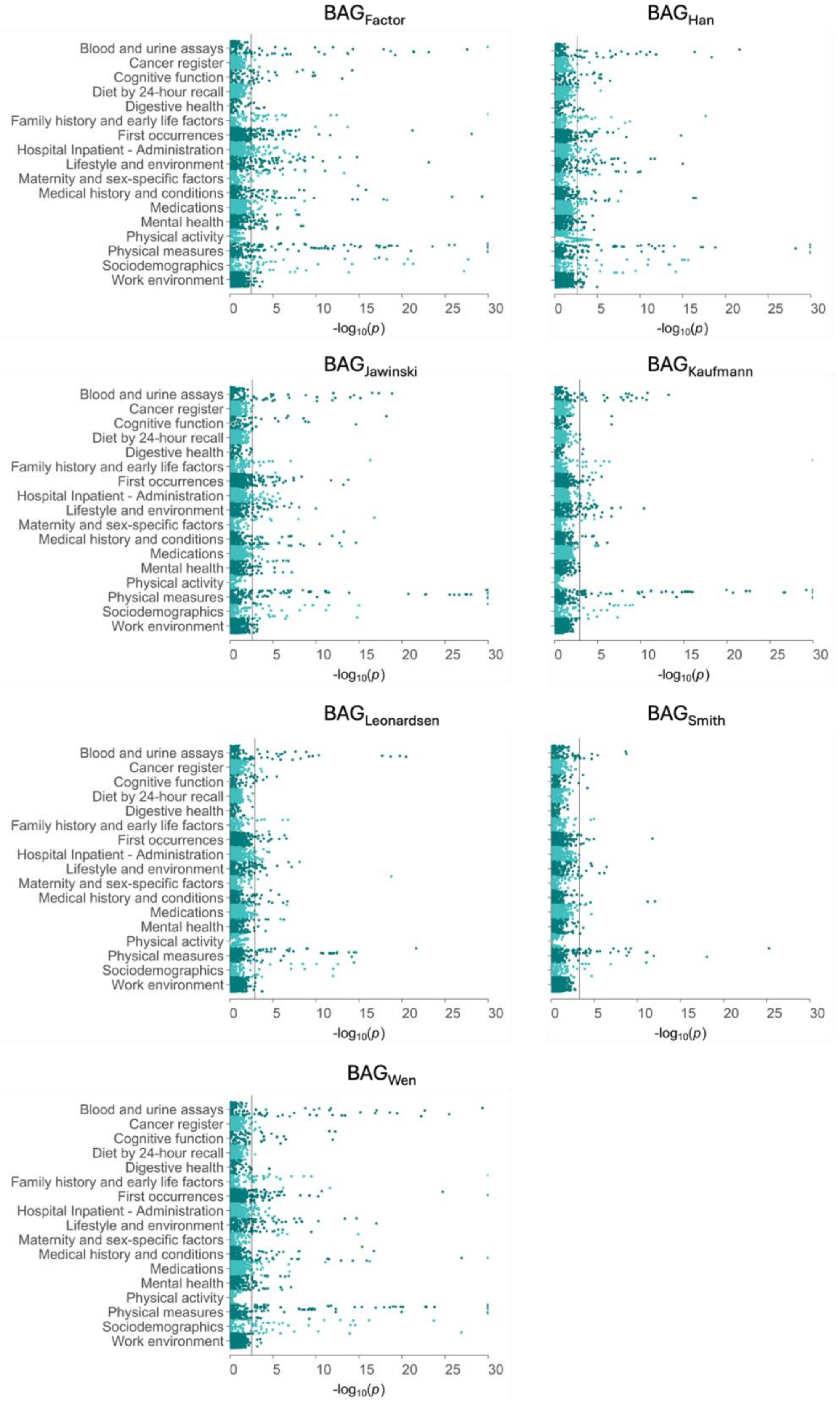

#### References


Akbarian, S., Liu, C., Knowles, J. A., Vaccarino, F. M., Farnham, P. J., Crawford, G. E., Jaffe, A. E., Pinto, D., Dracheva, S., Geschwind, D. H., Mill, J., Nairn, A. C., Abyzov, A., Pochareddy, S., Prabhakar, S., Weissman, S., Sullivan, P. F., State, M. W., Weng, Z., … Sestan, N. (2015). The PsychENCODE project. *Nature Neuroscience*, *18*(12), 1707–1712. https://doi.org/10.1038/nn.4156

Bashyam, V. M., Erus, G., Doshi, J., Habes, M., Nasrallah, I. M., Truelove-Hill, M., Srinivasan, D., Mamourian, L., Pomponio, R., Fan, Y., Launer, L. J., Masters, C. L., Maruff, P., Zhuo, C., Völzke, H., Johnson, S. C., Fripp, J., Koutsouleris, N., Satterthwaite, T. D., … Davatzikos, C. (2020). MRI signatures of brain age and disease over the lifespan based on a deep brain network and 14 468 individuals worldwide. *Brain*, *143*(7), 2312–2324. https://doi.org/10.1093/brain/awaa160

Bulik-Sullivan, B., Finucane, H. K., Anttila, V., Gusev, A., Day, F. R., Loh, P.-R., Duncan, L., Perry, J. R. B., Patterson, N., Robinson, E. B., Daly, M. J., Price, A. L., & Neale, B. M. (2015). An atlas of genetic correlations across human diseases and traits. *Nature Genetics*, *47*(11), Article 11. https://doi.org/10.1038/ng.3406

Bulik-Sullivan, B. K., Loh, P.-R., Finucane, H. K., Ripke, S., Yang, J., Patterson, N., Daly, M. J., Price, A. L., & Neale, B. M. (2015). LD Score regression distinguishes confounding from polygenicity in genome-wide association studies. *Nature Genetics*, *47*(3), Article 3. https://doi.org/10.1038/ng.3211

Buniello, A., MacArthur, J. A. L., Cerezo, M., Harris, L. W., Hayhurst, J., Malangone, C., McMahon, A., Morales, J., Mountjoy, E., Sollis, E., Suveges, D., Vrousgou, O., Whetzel, P. L., Amode, R., Guillen, J. A., Riat, H. S., Trevanion, S. J., Hall, P., Junkins, H., … Parkinson, H. (2019). The NHGRI-EBI GWAS Catalog of published genome-wide association studies, targeted arrays and summary statistics 2019. *Nucleic Acids Research*, *47*(D1), D1005–D1012. https://doi.org/10.1093/nar/gky1120

Chang, C. C., Chow, C. C., Tellier, L. C., Vattikuti, S., Purcell, S. M., & Lee, J. J. (2015). Second-generation PLINK: Rising to the challenge of larger and richer datasets. *Gigascience*, *4*(1), s13742-015-0047–0048. https://doi.org/10.1186/s13742-015-0047-8

Desikan, R. S., Ségonne, F., Fischl, B., Quinn, B. T., Dickerson, B. C., Blacker, D., Buckner, R. L., Dale, A. M., Maguire, R. P., Hyman, B. T., Albert, M. S., & Killiany, R. J. (2006). An automated labeling system for subdividing the human cerebral cortex on MRI scans into gyral based regions of interest. *NeuroImage*, *31*(3), 968–980. https://doi.org/10.1016/j.neuroimage.2006.01.021

Dörfel, R. P., Ozenne, B., Ganz, M., Alzheimer's Disease Neuroimaging Initiative (ADNI), Svensson, J. E., & Plavén-Sigray, P. (2025). Prediction of brain age using structural magnetic resonance imaging: A comparison of clinical validity of publicly available software packages. *medRxiv: The Preprint Server for Health Sciences*, 2025.03.13.25323902. https://doi.org/10.1101/2025.03.13.25323902

Endele, S., Nelkenbrecher, C., Bördlein, A., Schlickum, S., & Winterpacht, A. (2011). C4ORF48, a gene from the Wolf-Hirschhorn syndrome critical region, encodes a putative neuropeptide and is expressed during neocortex and cerebellar development. *Neurogenetics*, *12*(2), 155–163. https://doi.org/10.1007/s10048-011-0275-8

Evangelou, E., & Ioannidis, J. P. A. (2013). Meta-analysis methods for genome-wide association studies and beyond. *Nature Reviews Genetics*, *14*(6), 379–389. https://doi.org/10.1038/nrg3472

Feng, S., Liu, D., Zhan, X., Wing, M. K., & Abecasis, G. R. (2014). RAREMETAL: Fast and powerful meta-analysis for rare variants. *Bioinformatics (Oxford, England)*, *30*(19), 2828–2829. https://doi.org/10.1093/bioinformatics/btu367

Fischl, B. (2012). FreeSurfer. *NeuroImage*, *62*(2), 774–781. https://doi.org/10.1016/j.neuroimage.2012.01.021

Grasby, K. L., Jahanshad, N., Painter, J. N., Colodro-Conde, L., Bralten, J., Hibar, D. P., Lind, P. A., Pizzagalli, F., Ching, C. R. K., McMahon, M. A. B., Shatokhina, N., Zsembik, L. C. P., Thomopoulos, S. I., Zhu, A. H., Strike, L. T., Agartz, I., Alhusaini, S., Almeida, M. A. A., Alnæs, D., … Enhancing NeuroImaging Genetics through Meta-Analysis Consortium (ENIGMA)—Genetics working group. (2020). The genetic architecture of the human cerebral cortex. *Science (New York, N.Y.)*, *367*(6484), Article 6484. https://doi.org/10.1126/science.aay6690

Han, L. K. M., Dinga, R., Hahn, T., Ching, C. R. K., Eyler, L. T., Aftanas, L., Aghajani, M., Aleman, A., Baune, B. T., Berger, K., Brak, I., Filho, G. B., Carballedo, A., Connolly, C. G., Couvy-Duchesne, B., Cullen, K. R., Dannlowski, U., Davey, C. G., Dima, D., … Schmaal, L. (2021). Brain aging in major depressive disorder: Results from the ENIGMA major depressive disorder working group. *Molecular Psychiatry*, *26*(9), 5124–5139. https://doi.org/10.1038/s41380-020-0754-0

Jawinski, P., Forstbach, H., Kirsten, H., Beyer, F., Villringer, A., Witte, A. V., Scholz, M., Ripke, S., & Markett, S. (2025). Genome-wide analysis of brain age identifies 59 associated loci and unveils relationships with mental and physical health. *Nature Aging*, *5*(10), 2086–2103. https://doi.org/10.1038/s43587-025-00962-7

Kaufmann, T., van der Meer, D., Doan, N. T., Schwarz, E., Lund, M. J., Agartz, I., Alnæs, D., Barch, D. M., Baur-Streubel, R., Bertolino, A., Bettella, F., Beyer, M. K., Bøen, E., Borgwardt, S., Brandt, C. L., Buitelaar, J., Celius, E. G., Cervenka, S.,

Conzelmann, A., … Karolinska Schizophrenia Project (KaSP). (2019). Common brain disorders are associated with heritable patterns of apparent aging of the brain. *Nature Neuroscience*, *22*(10), 1617–1623. https://doi.org/10.1038/s41593-019-0471-7

Kavvoura, F. K., & Ioannidis, J. P. A. (2008). Methods for meta-analysis in genetic association studies: A review of their potential and pitfalls. *Human Genetics*, *123*(1), Article 1. https://doi.org/10.1007/s00439-007-0445-9

Kawaji, H., Kasukawa, T., Forrest, A., Carninci, P., & Hayashizaki, Y. (2017). The FANTOM5 collection, a data series underpinning mammalian transcriptome atlases in diverse cell types. *Scientific Data*, *4*(1), 170113. https://doi.org/10.1038/sdata.2017.113

Kharel, P., Singhal, N. K., Mahendran, T., West, N., Croos, B., Rana, J., Smith, L., Freeman, E., Chattopadhyay, A., McDonough, J., & Basu, S. (2023). NAT8L mRNA oxidation is linked to neurodegeneration in multiple sclerosis. *Cell Chemical Biology*, *30*(3), 308-320.e5. https://doi.org/10.1016/j.chembiol.2023.02.007

Leonardsen, E. H., Vidal-Piñeiro, D., Roe, J. M., Frei, O., Shadrin, A. A., Iakunchykova, O., De Lange, A.-M. G., Kaufmann, T., Taschler, B., Smith, S. M., Andreassen, O. A., Wolfers, T., Westlye, L. T., & Wang, Y. (2023). Genetic architecture of brain age and its causal relations with brain and mental disorders. *Molecular Psychiatry*, *28*(7), 3111–3120. https://doi.org/10.1038/s41380-023-02087-y

Liberzon, A., Subramanian, A., Pinchback, R., Thorvaldsdóttir, H., Tamayo, P., & Mesirov, J. P. (2011). Molecular signatures database (MSigDB) 3.0. *Bioinformatics (Oxford, England)*, *27*(12), Article 12. https://doi.org/10.1093/bioinformatics/btr260

Loh, P.-R., Tucker, G., Bulik-Sullivan, B. K., Vilhjálmsson, B. J., Finucane, H. K., Salem, R. M., Chasman, D. I., Ridker, P. M., Neale, B. M., Berger, B., Patterson, N., & Price, A.

L. (2015). Efficient Bayesian mixed-model analysis increases association power in large cohorts. *Nature Genetics*, *47*(3), 284–290. https://doi.org/10.1038/ng.3190

Mbatchou, J., Barnard, L., Backman, J., Marcketta, A., Kosmicki, J. A., Ziyatdinov, A., Benner, C., O'Dushlaine, C., Barber, M., Boutkov, B., Habegger, L., Ferreira, M., Baras, A., Reid, J., Abecasis, G., Maxwell, E., & Marchini, J. (2021). Computationally efficient whole-genome regression for quantitative and binary traits. *Nature Genetics*, *53*(7), 1097–1103. https://doi.org/10.1038/s41588-021-00870-7

Muñoz-Lasso, D. C., Romá-Mateo, C., Pallardó, F. V., & Gonzalez-Cabo, P. (2020). Much More Than a Scaffold: Cytoskeletal Proteins in Neurological Disorders. *Cells*, *9*(2), 358. https://doi.org/10.3390/cells9020358

Ramasamy, A., Trabzuni, D., Guelfi, S., Varghese, V., Smith, C., Walker, R., De, T., Coin, L., de Silva, R., Cookson, M. R., Singleton, A. B., Hardy, J., Ryten, M., & Weale, M. E. (2014). Genetic variability in the regulation of gene expression in ten regions of the human brain. *Nature Neuroscience*, *17*(10), Article 10. https://doi.org/10.1038/nn.3801

Schilling, S., Mehr, A., Ludewig, S., Stephan, J., Zimmermann, M., August, A., Strecker, P., Korte, M., Koo, E. H., Müller, U. C., Kins, S., & Eggert, S. (2017). APLP1 Is a Synaptic Cell Adhesion Molecule, Supporting Maintenance of Dendritic Spines and Basal Synaptic Transmission. *The Journal of Neuroscience*, *37*(21), 5345–5365. https://doi.org/10.1523/JNEUROSCI.1875-16.2017

Schmitt, A. D., Hu, M., Jung, I., Xu, Z., Qiu, Y., Tan, C. L., Li, Y., Lin, S., Lin, Y., Barr, C. L., & Ren, B. (2016). A Compendium of Chromatin Contact Maps Reveals Spatially Active Regions in the Human Genome. *Cell Reports*, *17*(8), 2042–2059. https://doi.org/10.1016/j.celrep.2016.10.061

Schulz, M.-A., Siegel, N. T., & Ritter, K. (2025). Brain-age models with lower age prediction accuracy have higher sensitivity for disease detection. *PLOS Biology*, *23*(10), e3003451. https://doi.org/10.1371/journal.pbio.3003451

Slenter, D. N., Kutmon, M., Hanspers, K., Riutta, A., Windsor, J., Nunes, N., Mélius, J., Cirillo, E., Coort, S. L., Digles, D., Ehrhart, F., Giesbertz, P., Kalafati, M., Martens, M., Miller, R., Nishida, K., Rieswijk, L., Waagmeester, A., Eijssen, L. M. T., … Willighagen, E. L. (2018). WikiPathways: A multifaceted pathway database bridging metabolomics to other omics research. *Nucleic Acids Research*, *46*(D1), D661–D667. https://doi.org/10.1093/nar/gkx1064

Smith, S. M., Elliott, L. T., Alfaro-Almagro, F., McCarthy, P., Nichols, T. E., Douaud, G., & Miller, K. L. (2020). Brain aging comprises many modes of structural and functional change with distinct genetic and biophysical associations. *eLife*, *9*, e52677. https://doi.org/10.7554/eLife.52677

Soumya Kumari, L. K., & Sundarrajan, R. (2024). A review on brain age prediction models. *Brain Research*, *1823*, 148668. https://doi.org/10.1016/j.brainres.2023.148668

Tafoya, L. C. R., Mameli, M., Miyashita, T., Guzowski, J. F., Valenzuela, C. F., & Wilson, M. C. (2006). Expression and Function of SNAP-25 as a Universal SNARE Component in GABAergic Neurons. *The Journal of Neuroscience*, *26*(30), 7826–7838. https://doi.org/10.1523/JNEUROSCI.1866-06.2006

The GTEx Consortium, Aguet, F., Anand, S., Ardlie, K. G., Gabriel, S., Getz, G. A., Graubert, A., Hadley, K., Handsaker, R. E., Huang, K. H., Kashin, S., Li, X., MacArthur, D. G., Meier, S. R., Nedzel, J. L., Nguyen, D. T., Segrè, A. V., Todres, E., Balliu, B., … Volpi, S. (2020). The GTEx Consortium atlas of genetic regulatory effects across human tissues. *Science*, *369*(6509), 1318–1330. https://doi.org/10.1126/science.aaz1776

Watanabe, K., Taskesen, E., van Bochoven, A., & Posthuma, D. (2017). Functional mapping and annotation of genetic associations with FUMA. *Nature Communications*, *8*(1), Article 1. https://doi.org/10.1038/s41467-017-01261-5

Wen, J., Tian, Y. E., Skampardoni, I., Yang, Z., Cui, Y., Anagnostakis, F., Mamourian, E., Zhao, B., Toga, A. W., Zalesky, A., & Davatzikos, C. (2024). The genetic architecture of biological age in nine human organ systems. *Nature Aging*, *4*(9), 1290–1307. https://doi.org/10.1038/s43587-024-00662-8

Zheng, Z., Liu, S., Sidorenko, J., Wang, Y., Lin, T., Yengo, L., Turley, P., Ani, A., Wang, R., Nolte, I. M., Snieder, H., LifeLines Cohort Study, Aguirre-Gamboa, R., Deelen, P., Franke, L., Kuivenhoven, J. A., Lopera Maya, E. A., Sanna, S., Swertz, M. A., … Zeng, J. (2024). Leveraging functional genomic annotations and genome coverage to improve polygenic prediction of complex traits within and between ancestries. *Nature Genetics*, *56*(5), 767–777. https://doi.org/10.1038/s41588-024-01704-y
